# An accurate genetic colocalization method for the HLA locus

**DOI:** 10.1101/2024.11.05.24316783

**Authors:** Guillaume Butler-Laporte, Tianyuan Lu, Sam Morris, Wenmin Zhang, Gavin Band, Fergus Hamilton, Amanda Chong, Kuang Lin, Ruth Nanjala, J Brent Richards, Mei-Hsuan Lee, Ling Yang, Pang Yao, Liming Li, Zhengming Chen, Yang Luo, Iona Y Milwood, Robin G Walters, Alexander J Mentzer

## Abstract

Genetic colocalization analyses are frequently conducted to determine if causal signals at a genetic locus are shared between two phenotypes. However, colocalization is rarely undertaken at the HLA locus, due to its complex linkage disequilibrium (LD) and high polymorphism density. This lack of genetic causal inference method limits our ability to translate HLA associations into therapeutic targets. Here we present a method that uses HLA alleles, instead of nucleotide variants, to perform genetic colocalization of two traits at HLA genes. The method, which we call HLA-colocalization, works by controlling for LD using a Bayesian variable selection algorithm (here implemented with SuSiE), then performing Bayesian regression on the resulting posterior inclusion probabilities. We first show through simulation that the method correctly identifies truly colocalizing genes. We then test the method in two positive control scenarios, showing colocalization between hepatitis B and liver disease at *HLA-DPB1*, and between Epstein-Barr virus and multiple sclerosis at *HLA-DRB1* and *HLA-DQB1*. Lastly, we perform a large colocalization scan between multiple viruses and auto-immune diseases, demonstrating that the method is well calibrated, and uncovering multiple biologically plausible novel causal associations, such as cytomegalovirus and ulcerative colitis. To our knowledge, HLA-colocalization is the first accurate genetic colocalization method for the HLA locus (github: https://github.com/DrGBL/hlacoloc).

## Introduction

The human leukocyte antigen (HLA) cluster of genes on chromosome 6 of the human genome is associated with multiple autoimmune, inflammatory, and infectious conditions^1–3^. It contains genes that are critical for a functioning innate and adaptive immune response including those encoding complement proteins, as well as class I and II HLA proteins that are responsible for presenting self and foreign peptide to CD8+ and CD4+ cells respectively^4^. It is widely recognised as one of the most complex genetic loci in the human genome, due to its high density of structural and single nucleotide polymorphisms, the complex long-range linkage disequilibrium^2^ (LD), and the fact that multiple independent associations may be observed across the locus with single traits.

These genetic complexities, that are unique to HLA, prohibit the application of genetic epidemiological causal inference methods, such as Mendelian randomization or genetic colocalization, that have resulted in significant translational breakthroughs and new therapeutic discoveries in other regions of the genome^5,6^. In the case of Mendelian randomization, the HLA locus likely breaks the core assumption of absence of horizontal pleiotropy (i.e. the HLA locus is associated with too many traits or diseases for any HLA SNP instrument to confidently be only associated with an outcome through its role on the exposure). In the case of genetic colocalization, the long-range LD is either computationally intractable (i.e. the algorithms do not converge when including classical variants such as single nucleotide polymorphisms, SNPs), or the outputs provide biologically uninformative results even when colocalization is probable (i.e. it cannot identify specific HLA or loci that drive the colocalization). That is, even if genetic colocalization is observed at the HLA, it is still difficult with currently available methods to pinpoint specific genes or alleles within the HLA that explain the observed shared genetic signal between two phenotypes. Hence, given the breadth of diseases linked with HLA and the potential for translational opportunity, a method that could perform genetic colocalization and inform biologically causal components of the HLA is a great unmet need.

In what follows, we present an overview of our proposal of the underlying architecture of HLA gene and allele associations with disease traits. We then outline a method that exploits this model, and tests for colocalization at HLA genes between two traits, thus finding potential links between those tested phenotypes. This method only requires the key assumptions that the causal HLA genes (there may be more than one) for the traits must be included in the analysis, and that traits are analyzed in populations with the same LD structure. Moreover, colocalization results are given at the level of genes, rather than group of SNPs (e.g. the probability of hepatitis B and liver cirrhosis colocalizing at *HLA-DPB1* is 99%).

We test the method using simulations in cohorts of diverse genetic ancestries derived from the UK Biobank^7^, then using known positive control scenarios, we show results of colocalization at varying number of HLA allele fields to show that these can provide biologically relevant insight into the HLA. Specifically, we show how Epstein-Barr virus seropositivity colocalizes at the HLA with multiple sclerosis in European ancestry populations, and how hepatitis B antigen positivity colocalizes with liver disease in the East Asian populations^8^. Lastly, we perform a large-scale HLA-colocalization analyses of pathogen serology and autoimmune diseases, finding novel colocalizing genetic signals, opening up potentially unexplored links between pathogens and disease.

## Results

### A theoretical architecture of HLA-disease associations; the gene-allele signature

Other less complex regions of the genome have genetic associations with disease observed as a result of causal, predominantly biallelic, SNPs affecting gene transcription or their gene product function, with surrounding SNPs associated through LD (**Fig. 1a**). In contrast, associations observed in the HLA region typically show many other SNPs apparently associated as result of the long-range LD^9–12^ in addition to those in local LD. For most traits with SNP associations across the HLA, our current understanding is that the associations are a result of multiple independent associations between classical HLA gene alleles, typically focussing on the class I (*HLA-A*, -*B*, and –*C*) and class II (*HLA-DR*, -*DQ* and -*DP* heterodimer) genes^9–12^ (although notable exceptions exist^13^).

**Figure 1:**
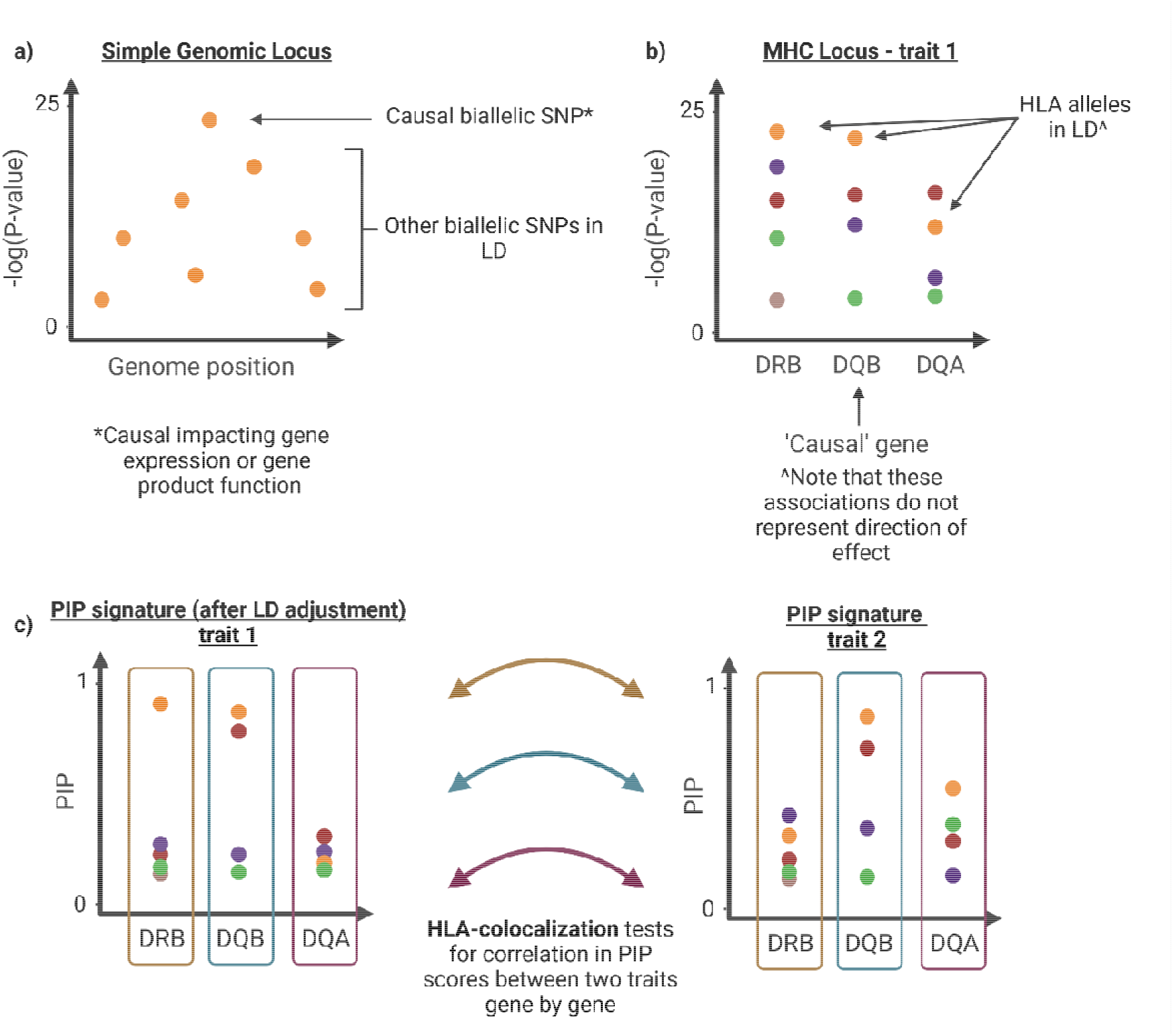
Visual representation of LD at the HLA and HLA-colocalization. **a)** Distribution of effect sizes in a typical SNP-based association study. P-values decay as variants become further away from the lead SNP. This is also observed at the HLA locus when a SNP association is observed with a trait, and the LD may span the entirety of the HLA locus. **b)** In contrast to SNP associations, HLA allele associations do not display such rapid decaying LD with increasing genomic distance. This is because HLA alleles for a given gene all share the same position. However, between gene LD still exists, and is represented by the matching colours in the figure. **c)** After using BVS model, we obtain the most predictive HLA allele combination for both the trait 1, and a comparator trait 2. In some cases, only alleles at one gene will be predictive (as for the red dots in trait 1). In other cases, alleles from the same haplotype may appear predictive at more gene loci (e.g. yellow dots, with alleles at 2 genes in trait 1). In most cases, no HLA alleles will be predictive of the trait above and beyond the other more predictive alleles (dots of other colours). This significantly reduces the problem of LD in colocalization. HLA-colocalization tests for correlation in PIP signatures gene-by-gene, to define the gene with the highest degree of correlation between two traits. In this example HLA-DQB(1) will exhibit the greatest level of correlation.

In what follows we refer to HLA alleles using the standard nomenclature, which consists of the gene name, followed by 4 colon-separated fields that provide information on serotype, protein altering variants, synonymous variant, and non-coding variants respectively (e.g. allele HLA-A*01:01:01:01 is a classic example of a 4-field allele). This nomenclature is used due to the high number of polymorphisms at HLA genes. Each field describes a set of genetic variants that together represent a given version of the HLA gene (i.e. an allele) to a certain definition. Depending on the technology used for genotyping, HLA alleles can be described to any given field length, with increasing resolution of underlying single variants characterised as the number of fields increases. Thus, this allele nomenclature inherently describes clusters of variants forming the functional HLA molecule.

Upon imputation, or sequencing, of HLA alleles and testing of the resultant allele associations with disease traits, multiple alleles in many HLA genes have observed associations. Several alleles in different genes frequently have near-equivalent association test statistics owing to LD (**Fig. 1b**). Differentiating causal alleles within genes, assuming a similar architecture to less complex loci, has near-ubiquitously been elusive. For example, HLA haplotypes *DR1, DR2, DR3*, and *DR4* are all strongly associated with the risk of type 1 diabetes mellitus, but span multiple class II HLA genes (most significantly *HLA-DRB1* and *HLA-DQB1*)^14^.

Another unique observation with HLA associations is that not only are there single alleles in significant association with disease traits in each gene, but many other alleles in each gene also demonstrate associations with the trait^2,15^. The direction of effect of these alleles on the trait may be positive (risk increasing in the case of binary disease) or negative (protective). The explanation for these observations can be postulated to be a result of HLA alleles representing single-unit proteins that bind and present relevant self-(class I) or foreign-(class II) peptides in either shared or distinct ways^16^. Those alleles within genes with shared properties often have shared peptide-contacting amino acid residues, whereas other amino acids at those positions may explain opposing effects. Together, multiple alleles within a gene represent a spectrum of potential effects on the trait depending on their ability to bind and present peptide. However, the measured effect (and resultant association statistic) of any one allele will be a combination of the true effect on the trait, and LD with any other allele in another gene that may influence that same trait. We propose that if we can define the alleles within each gene that are likely to be most predictive of any trait, after adjusting for complex LD, we may be able to define a ‘signature’ of association for each HLA gene, that may then be tested with other traits to find the probability of colocalization (**Fig. 1c**).

### Overview of the HLA colocalization method

Here, we present HLA-colocalization, an easy-to-use Bayesian method that allows for the assessment of genetic colocalization of two traits at HLA genes using summary statistics through the generation of LD-adjusted allelic signatures of association. Compared to standard genetic colocalization^17,18^ methods, this method does not colocalize at the level of biallelic SNPs, but rather at the level of whole HLA genes using HLA allele nomenclature described above. The method defines HLA alleles as multiallelic variants at any given *HLA* gene. Hence, HLA allele based colocalization seeks to find which genes, rather than which SNP, harbor the shared genetic determinants for a given pair of traits. However, similar to SNP-based colocalization, HLA colocalization also tests the property that allele true effect sizes are proportional between the two traits at the causal HLA gene. The key difference being that in SNP-based colocalization, proportionality is assumed at one variant and observed through LD at the entire locus, whereas at HLA alleles the proportionality property is intrinsic to a gene and gets obscured by LD (rather than reinforced). To avoid ambiguity, in the remainder of the text, we will use the term “allele” to refer exclusively to HLA alleles as described above, and we will use “SNP” to refer to single-nucleotide variants.

Modern SNP-based colocalization methods vary, but most of them generally work in two steps. In the first step, sets of largely independent SNPs are identified. These sets are deemed to be the most likely determinant of their respective phenotypes and are determined through different algorithms accounting for LD such as conditional analyses^6^ or Bayesian variant selection^17^ (BVS). In the second step, algorithms determine if the sets of SNPs selected for each phenotype in the first step are shared between those phenotypes. Measuring how much is shared between these sets of variants is also done in varying ways such as multiplying posterior inclusion probabilities (PIPs) or Bayes factors, for example^6,19,20^.

HLA-colocalization follows the same general approach. In the first step, we select a set of HLA alleles which are most predictive of each trait. This is done with a BVS algorithm (SuSiE^19^), resulting in each HLA gene being assigned a set of alleles with varying PIPs. Alleles with high PIPs are interpreted as being more predictive of the phenotype at that gene. Working with HLA alleles allows for the simplification of the LD and makes the BVS algorithm robust to the HLA LD structure. This distribution of PIPs then provides a causality signature for each gene that we use in the second step, where we measure how similar these causality signatures are for each gene between traits. Phenotypes which share a gene with similar causality signature are said to colocalize at that gene. In our HLA-colocalization method these steps are performed using Bayesian methods, allowing for a final probability of colocalization at each HLA gene. Specifically, if two phenotypes have a high probability of HLA colocalization at the same gene, then they are likely to share the same genetic determinant at that gene. Hence, if one assumes that the HLA locus is causal for the phenotypes, then the mechanism behind this causality is shared between the two phenotypes at the gene(s) with high probability of colocalization. We again emphasize that the second step is done independently for each gene, and that one final probability of colocalization is provided for each gene. Note that similar to SNP-based colocalization, direction of causality from one phenotype to the next is neither tested nor assumed. However, in contrast to SNP-based colocalization, this method provides a probability that two phenotypes colocalize at an HLA gene, rather than a locus.

HLA-colocalization handles the two main problems with SNP-based colocalization at the HLA described in the introduction. First, it alleviates LD bias enough that BVS becomes reliable. That is, while there is still considerable LD between some HLA alleles at different genes (**Fig. 1b**), there is by definition no LD between alleles of the same gene (the probability of carrying any given two HLA allele at a certain gene depends only on populational allele frequency). This considerably simplifies LD at the HLA and allows BVS to efficiently select the most predictive alleles in the first step of the algorithm (**Fig. 1c**). Second, by working with HLA alleles directly, we introduce biological context to colocalization, since the result can be directly interpreted at the level of individual HLA genes.

### Simulation

We used simulation of two quantitative traits to determine the PIP estimates expected if there was a true colocalization between two traits at one or more HLA gene using our method, compared to estimates expected if there was no colocalization, whilst varying the proportion of variance explained by the HLA genes on the likelihood of both traits. To do this we ran 50,000 simulations (10,000 per ancestries) of random pairs of traits using 3-field HLA allele calls obtained from whole-exome sequence (WES) data available from UK Biobank (UKB)^2^. For those simulations defining a true colocalization, the causal genes were randomly selected with the proportionality factor for each allele within that gene randomly assigned to both traits. The final proportion of variance explained by these alleles and genes was then averaged by adding random error, and linear regression was performed assuming an additive model. These simulations are designed to capture the model outlined above, i.e. where multiple alleles at a single gene may affect the trait with a spectrum of effect sizes, such that colocalizing traits have proportional effect sizes (which are on the logistic scale in our binary trait simulations).

Simulation results are summarized in **Fig. 2**. As the variance explained by HLA genes increased, the colocalization probability increased rapidly for truly colocalizing genes, and remained low for non-colocalizing genes (**Fig. 2a**). Importantly, this was observed in all continental ancestries, despite differences in LD architecture and sample size (2,647 in east Asians, 3,101 in admixed Americans, 8,734 in Europeans, 9,388 in Africans, and 9,449 in south Asians). The probability of colocalization were also well calibrated, in that for any N, around N% of genes with a probability of N% were truly colocalizing (**Fig. 2b**). Reassuringly, while the method was less well calibrated in cases where the genes explained a lower proportion of the trait’s variance, this was erred on side of giving a lower probability. We hypothesized that this was probably an issue with SuSiE which was not able to assign high PIPs to genes with small R2. Indeed, when we restricted the analysis to genes where at least one allele had a PIP>50%, the calibration was almost perfect (**Fig. 2c**). Assessing the method’s ability to differentiate between colocalizing and non-colocalizing genes, the area under the receiver operating characteristic curve increased from an average of 60.7% in HLA genes simulated to explain 0-3% of a trait’s variance, to an average of 89.7% in HLA genes explaining 6-9% of a trait’s variance (**Fig. 2d**, see **Supp. Fig. 1** for AUCs values by ancestries). We note that our simulations were deliberately conservative, as the HLA is known to explain a much higher percentage of certain traits’ variance (e.g. 42.8% in type I diabetes mellitus^21^).

**Figure 2.**
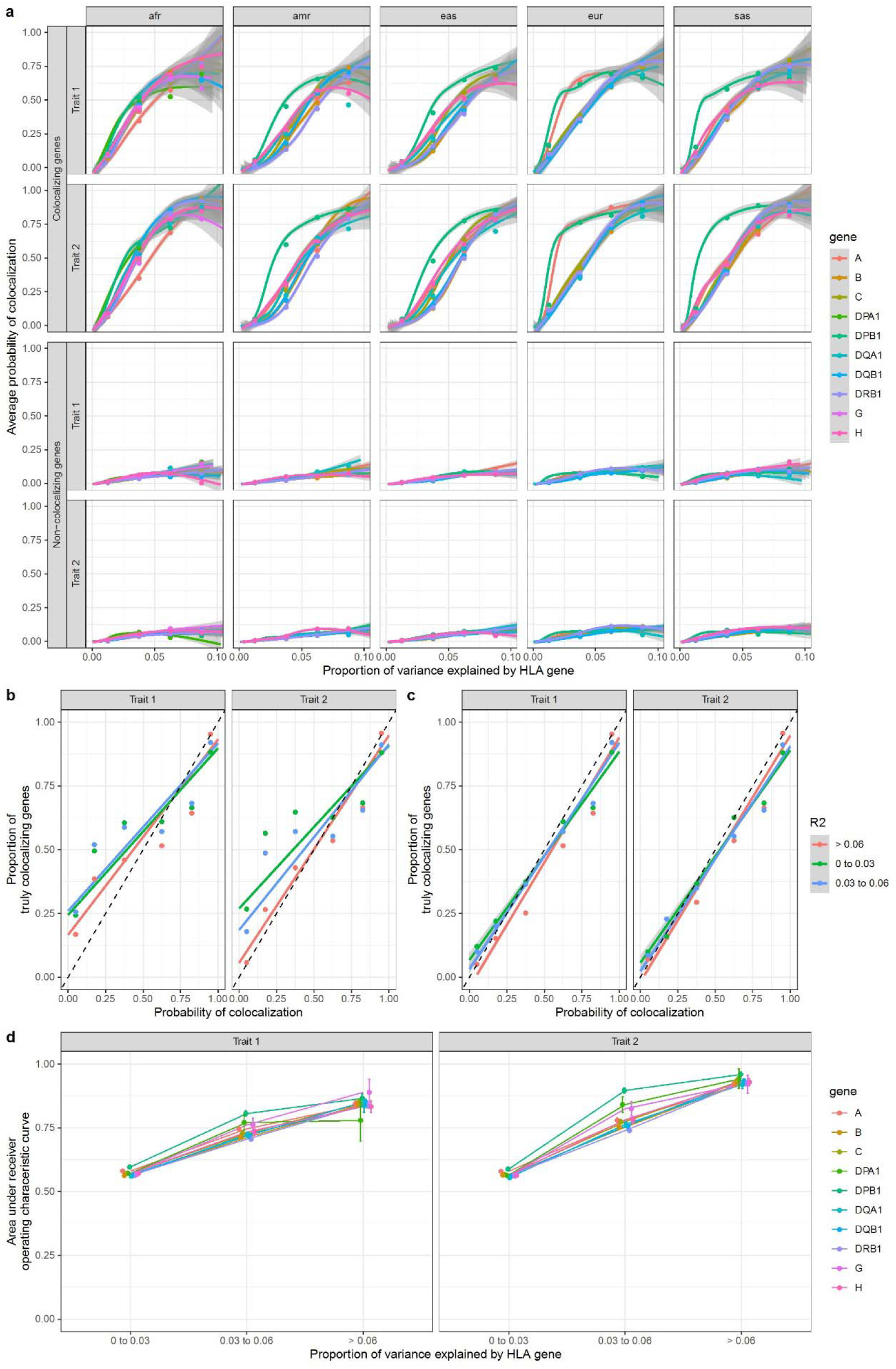
HLA allele HLA-colocalization simulation results for quantitative traits. Pairs of quantitative traits were simulated having either true overlap, or no true overlap between causal HLA alleles, using a bivariate normal model as described in Methods. In each simulation a total proportion of trait variance explained was assumed. A total of 50,000 simulations (10,000 per ancestry group) were performed covering different parameter values (*Methods*). HLA allele distributions were simulated using UK Biobank participants. **a)** The average posterior probability of colocalization in truly colocalizing increases with the amount of phenotype variance explained by each gene, as expected. The average posterior probability of colocalization in truly non-colocalizing genes remains stable with increasing variance explained. The lines were drawn using a generalized additive model with *geom_smooth* in R. The grey area represents 95% confidence intervals. The individual dots represent the average in the corresponding variance bins. **b)** The proportion of simulated genes that were truly colocalizing shown as a function of the probability of colocalization. This is close to the identity line, though errs on the more conservative side for genes with lower R2. **c)** The deviation from the identity line is largely due to situations where SuSiE is unable to assign a PIP larger than 50% in at least one allele at a gene. When we restrict to genes with a minimal PIP of 50%, the method is almost perfectly calibrated. **d)** Average area under the curve as a function of variance explained for each gene. For this plot, average ROC area under the curve across ancestry was shown. Legend: afr: African genetic ancestry, amr: Admixed American genetic ancestry, eas: East Asian genetic ancestry, eur: European genetic ancestry, sas: South Asian genetic ancestry.

We note that as with regular SNP-based colocalization, HLA-colocalization works only if there is a sufficient amount of genetic variation affecting the trait. Indeed, in our simulation, we only considered genes with 10 or more alleles.

We then looked at falsely colocalizing genes. To do this, we calculated the proportion of non-colocalizing genes with a probability of colocalization above 80% under varying circumstances. As expected, the proportion was higher in simulations where less genes were truly colocalizing then in simulations with more (**Supp. Fig 2a**), since more truly colocalizing genes decreases the chance of a random false colocalization. It also happened more often for genes which explained a larger portion of the traits, since genes with lower R2 are less likely to colocalize in the first place (**Supp. Fig 2c**). However, we found that the proportion was slightly higher in Europeans (1.6%) compared to other ancestries (e.g. admixed Americans at 0.58%) (**Supp. Fig 2d**). We also found that the proportion was higher at *HLA-DRB1* (1.45%) and *HLA-C* (1.32%) than other genes (e.g. *HLA-DQA1* 0.84%) (**Supp. Fig 2b**). Nevertheless, overall, only 1.1% of non-colocalizing genes were given a probability of colocalization above 80%.

Lastly, we performed a similar simulation for two binary traits (**Methods**) and obtained similar results (**Supp. Fig. 3-4**).

### Hepatitis B virus and liver diseases HLA-colocalization

We next applied our colocalization method to investigate the shared genetic architecture of measured human antibody responses against hepatitis B virus (HBV), and liver disease (including cancer). This was done in the China-Kadoorie Biobank (CKB), with HLA alleles imputation done at the G-group resolution on the HLA Michigan Imputation Server. We considered this analysis as a positive control since in East Asian populations the most cause of liver disease is chronic hepatitis B infection^22^ and thus we would expect a significant sharing of genetic architecture. There is strong evidence that immunity to HBV, thus influencing risk of chronic infection and sequelae, is in part determined by HLA variants, specifically at *HLA-DPB1*^*23*^. HLA association studies were performed on hepatitis B surface antigenemia (cases: 3,097, controls: 97,543), and on liver disease or liver cancer (case: 3,325, control: 97,315). Our HLA-colocalization method found that the expected gene colocalizes for the two traits (*HLA-DPB1* colocalization probability of 100%). It also provided weak support for colocalization at *HLA-DRB1* (P = 31%) and *HLA-DQB1* (P = 29%). These results are summarized in **Fig. 3**, which shows the original betas from HLA allele association studies in part *a*, and the resulting PIPs obtained from SuSiE in part *b* (see also **Supp. Data 1**).

**Figure 3:**
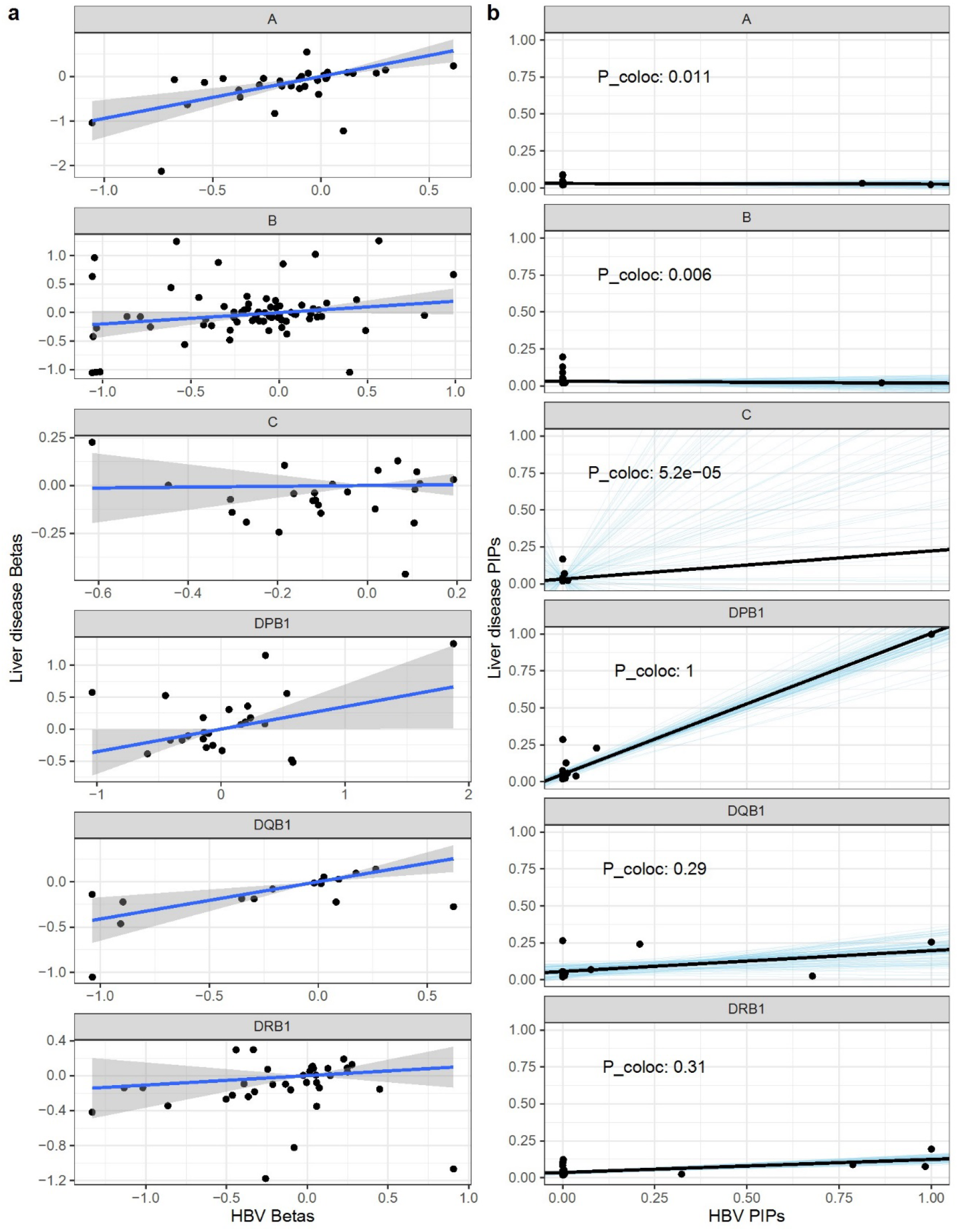
Liver disease and HBV antigenemia HLA-colocalization. **a)** linear regression (with 95% confidence intervals) of beta coefficients from the additive HLA allele association studies. *b)* Bayesian regression of HBV and liver disease PIP causal signature. The black lines show the egression fit, while the blue lines show 100 random draws from the posterior distributions. The resulting probabilities of HLA-colocalization (P_coloc) are also written for ease. Hence, after Bayesian variable selection at the HLA locus, *HLA-DPB1* shows evidence of shared liver disease and HBV genetic architecture.

Lastly, given that the above analysis was done in the same sample for HBV and liver disease phenotypes, we performed an analysis using data from a HLA association study of HBV infection in an east Asian genetic ancestry cohort from the Taiwan Biobank^24^. For this analysis, HLA allele imputation was done using HIBAG for class II genes only. For HLA-colocalization, analyses were limited to *HLA-DRB1, HLA-DQB1*, and *HLA-DPB1*, as *HLA-DPA1* and *HLA-DQA1* did not have enough alleles for the algorithm to converge. LD measures (r) between HLA alleles were taken from the CKB cohort. As expected, *HLA-DPB1* colocalized with a probability of 100%, while other genes did not show evidence of colocalization (**Supp. Fig. 5**).

### HLA-colocalization of Epstein-Barr virus antibody and multiple sclerosis risk

We next applied our colocalization method human antibody responses against Epstein-Barr virus (EBV), with multiple sclerosis disease (MS) risk. EBV and MS have long been reported to be associated, with a recent large-scale prospective cohort showing a clear temporal association between the two traits, with most cases of MS being preceded by EBV. In genetic studies, the association between *HLA-DRB1*\*15:01 and both MS and EBV antibody levels has been observed in multiple independent cohorts of different ancestries^1,3,25–27^. Similarly, *HLA-DQB1*\*02:01 has been linked to MS and EBV in Europeans^1,3,28^ but is in LD with *HLA-DRB1*03:01*.

We used a subset of individuals from UKB with serological measurements measured against two EBV antigens^1,3^, and using their associated whole-exome sequencing 3-field resolution HLA allele calls, we performed HLA-colocalization with a case control HLA-allele analysis of multiple sclerosis risk, again using individuals from UKB. We ran additive model HLA allele association studies on levels of inverse quantile normalized viral capsid antigen (VCA, n = 7,741) and EBV nuclear antigen-1 (EBNA1, n = 7247) antibodies, and on multiple sclerosis (cases = 2,363, controls = 427,459).

**Fig. 4** shows the results comparing the frequentist regression of distributions of betas of HLA allele associations with each trait, using VCA antibody response, alongside the results of the Bayesian HLA-colocalization for the same traits. This demonstrates firstly that where linear regression of betas may suggest a correlation between MS risk and VCA antibody response shared at either HLA-DQB1 or DRB1, the Bayesian HLA colocalization method supports previously reported associations between exposure to EBV (as measured by VCA levels) and multiple sclerosis risk being genetically linked at *HLA-DRB1* (P = 96%). Equivalent results were obtained for EBNA1 antibody levels and MS risk (*HLA-DRB1* P = 86%), but with additional support for *HLA-DQB1* (P = 100%) (**Supp. Fig. 6, Supp. Data 1**).

**Figure 4:**
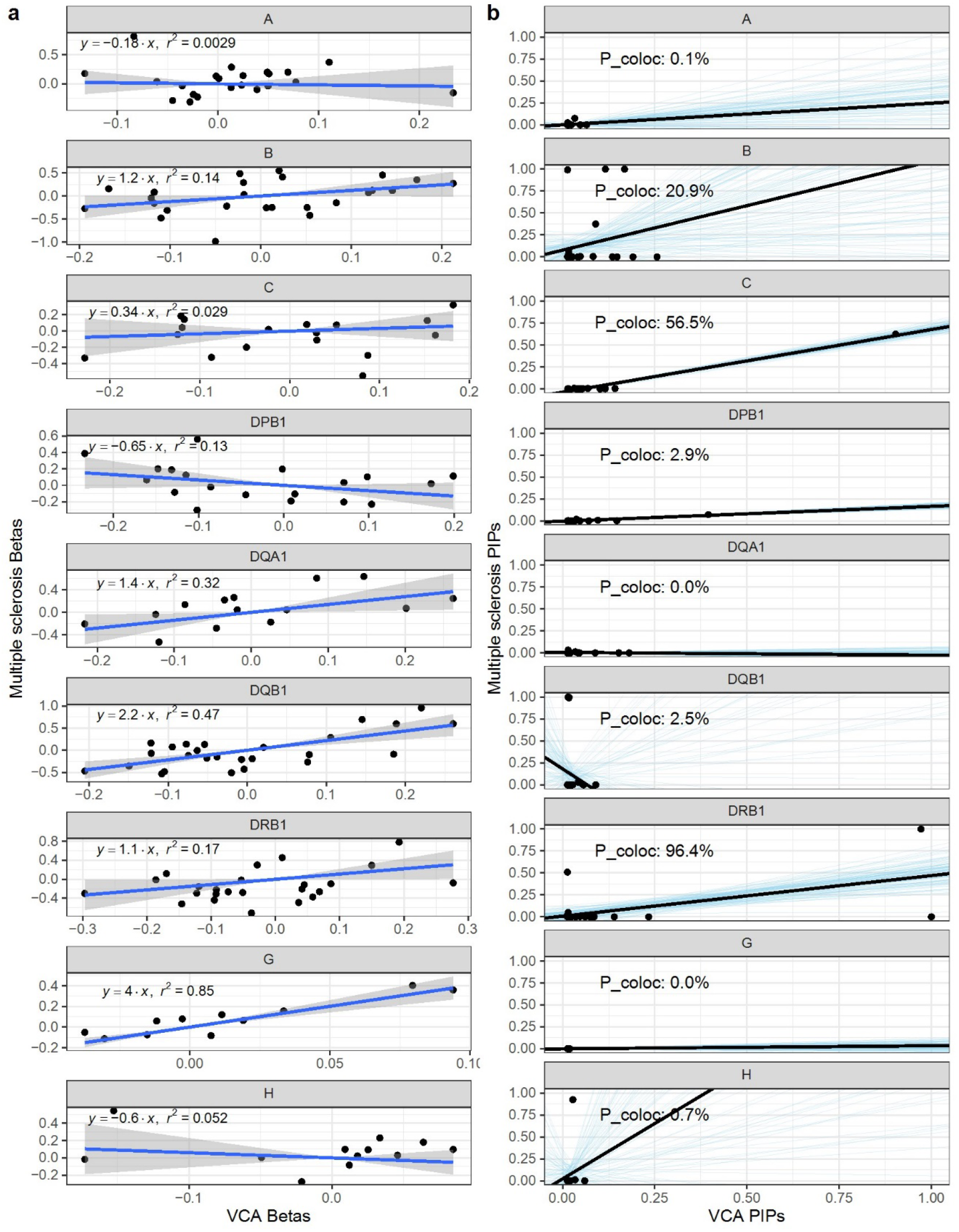
VCA and Multiple sclerosis HLA-colocalization. **a)** linear regression (with 95% confidence intervals) of beta coefficients from the additive HLA allele association studies. **b)** Bayesian regression of multiple sclerosis and VCA PIP causal signature. The black lines show the regression fit, while the blue lines show 100 random draws from the posterior distributions. The resulting probabilities of HLA-colocalization (P_coloc) are also written for ease. Hence, after Bayesian variable selection at the HLA locus, both *HLA-DQB1* and *HLA-DRB1* show evidence of shared multiple sclerosis and VCA genetic architecture.

The EBV and MS analysis above used a partially overlapping cohort of participants in the UK Biobank. However, in practice, colocalization is often performed in independent cohorts using summary statistics and an LD reference panel. We therefore repeated the analysis, but this time using a large independent cohort of MS cases (n=17,465) and controls (n=30,385) from the International Multiple Sclerosis Genetics Consortium (IMGSC) instead of participants with MS in the UK Biobank. The LD reference panel was obtained from European genetic ancestry UK Biobank but excluding participants with measured EBV antibody levels. Hence, summary statistics from the two phenotypes and the HLA allele LD reference panel were fully independent. Note that for this analysis, summary statistics were only available for the *HLA-A, HLA-B, HLA-C, HLA-DQB1*, and *HLA-DRB1*. Again, we found his probability of colocalization at *HLA-DRB1* for VCA (P = 85%, **Supp. Fig. 7**). However, for EBNA, colocalization probabilities decreased to 10% for *HLA-DRB1* and to 7% for *HLA-DQB1* (**Supp. Fig. 8, Supp. Data 1**). Together, these results strongly support a link through *HLA-DRB1* between EBV exposure and MS risk. Further, while using a full two-sample approach likely leads to some loss of power, the method still performs well in this scenario.

### Human infection antibody responses and auto-immune disease risk

Lastly, to measure the performance of our method and find potentially novel colocalizing associations on a larger scale, we performed HLA-colocalization on the HLA-wide association analyses of all infection antibody levels available in UKB, compared against HLA associations with 10 auto-immune diseases with well-described strong causal signals identified at the HLA^2^: asthma, multiple sclerosis, polymyalgia rheumatica and giant cell arteritis (PMR-GCA), rheumatoid arthritis, psoriasis, ankylosing spondylitis, auto-immune thyroid disorders, type 1 diabetes mellitus (T1D), Coeliac disease, and ulcerative colitis. The selected infectious agents were all viruses: cytomegalovirus (CMV), EBV, JC virus (JCV), Merkel cell polyomavirus (MCV), and varicella zoster virus (VZV). As expected, the majority of pairs of traits did not colocalize at any tested HLA gene. Only 6.3% of tested pairs of traits showed HLA-colocalization probability higher than 90%. Furthermore, 88.9% of pairs showed a probability of HLA-colocalization of less than 30% (**Supp. Fig. 9**). These suggest that the method is well calibrated to real-world data.

Of the pertinent high probability colocalizing pairs of traits, we find that EBV (measured with EBNA serology) colocalizes at the HLA with many auto-immune diseases: T1D at *HLA-DRB1* (P = 100%), auto-immune thyroid disorders at *HLA-DPB1* (P > 99%), asthma at *HLA-DQB1* (P > 99%), and PMR-GCA at *HLA-DQB1* (P > 99%). EBV has been tentatively linked to be part of the pathophysiology of most of these diseases^29,30^. We also observed colocalization between demyelinating disease and two polyomaviridae: JCV and MCV both at *HLA-DRB1* (P > 99%). JCV is a known cause of demyelinating diseases such as progressive multifocal leukoencephalopathy^31^, whereas MCV has been linked with the development of chronic inflammatory demyelinating polyneuropathy^32^, though this colocalization could also reflect the similarity between the two polyomaviridae. Interestingly, we found that CMV colocalizes strongly (using both the pp52 and pp150 antigens) with ulcerative colitis at *HLA-DRB1* (P > 99%). CMV is known to be one of the most common complications of ulcerative colitis and its immunosuppressive therapy^33–35^, but is also hypothesized to be involved in the pathogenesis of the disease and the severity of its acute flares^36^. Hence, the results from HLA-colocalization matches what can be observed in clinical practice.

Of the class I HLA genes, the strongest signals were found for VZV, which colocalized at *HLA-B* (P > 98%) with multiple auto-immune diseases: T1D, PMR-GCA, rheumatoid arthritis, multiple sclerosis or demyelinating diseases, Coeliac disease, and asthma. VZV is also suspected to be involved in many of these diseases, though more research is needed to understand the direction of causality. See **Supp. Data 2** for the full results.

### On the importance of using the correct HLA allele LD reference panel

As a last test for our algorithm, we wanted to determine how robust it was to a mismatch between the cohort used for the HLA allele association studies, and the cohort used in for the HLA allele LD reference. For this, we performed the same analysis as above on HBV and liver cirrhosis in the CKB cohort. However, we changed the LD reference used to perform the SuSiE step for HBV infection, using the non-east-Asian ancestries in the UKB (same as for our simulations above). The probability of colocalization at *HLA-DPB1* dropped from 100% in the correct analysis in east-Asians, to 99.9% in Africans, 85.9% in Europeans, 82.3% in south Asians, and 7.5% in admixed Americans. This confirms our suspicion that, like SNP based colocalization, the choice of LD reference panel is critical to ensure valid results.

## Discussion

Genetic colocalization methods are a useful causal inference tool which has been successfully applied to many loci across the genome. However, usual SNP-based methods fail at the HLA due to its complex LD and high polymorphism density. This has limited opportunities to translate genetic findings at the HLA locus into actionable therapeutic targets. Here, we have presented a genetic colocalization method which provides an accurate measurement of the degree of genetic architecture shared between two traits at HLA genes. Simulations and real-world application to two well established pairs of human diseases demonstrated high accuracy and low false positive signal rate. Lastly, a large-scale screen of colocalization between viral serologies and autoimmune diseases demonstrated that the method was well-calibrated, and still able to discover novel associations with biological and clinical plausibility (e.g. CMV and ulcerative colitis^33–35^).

However, there are still important caveats to HLA-colocalization. Most of these are similar to those encountered in SNP-based genetic colocalization. First, HLA-colocalization requires that sufficient genetic variation is captured by the HLA alleles. In our simulation, the BVS algorithm would often fail to converge for genes with less than 10 alleles. This fits with the intuition that the more information is given about LD architecture at a locus (by expanding the LD matrix), the easier it is to recover the most informative alleles for each trait. Hence HLA-colocalization can only be used in cohorts with enough genetic diversity at the HLA. In practice this also means that the cohort needs to be large enough. While what constitutes large enough depends on the trait, the cohort, and the effect size, it is clear that the method can only work if SuSiE is able to assign a high PIP to at least one HLA allele. Pragmatically, this means that our method is likely only expected to perform well if HLA allele association studies are able to identify at least one allele with a genome-wide significant p-value (< 5×10^-8^), which is also the minimum value we used in our simulations.

Second, our method also assumes that at least one of the HLA genes is causal for the trait. This is similar to the SNP-based colocalization assumption that there be at least 1 causal SNP at the locus for each phenotype. In the case of HLA allele colocalization, this means that the analysis needs to include all genes for which there could be a causal allele. This also implies that HLA-colocalization at an HLA gene does not provide information on whether the shared causal effect is due to coding variants, or due to non-coding variants that tag the relevant HLA alleles. It also means that lack of colocalization at a gene does not imply that this gene is not causal for the traits. Indeed, it could be causal for one or even both traits (just not in a colocalizing way), or that there was not enough statistical power. Nevertheless, any resultant probability suggesting colocalization can at least prioritise the locus for downstream translational or functional studies and can be used to support that the colocalizing gene is causal for both traits. For example, our results add further support that a vaccine preventing EBV infection could potentially prevent multiple sclerosis, and that prioritising DRB1 presented peptides could be advantageous.

Lastly, HLA colocalization requires an LD matrix between HLA alleles which can come from a reference population. If this LD matrix is not available owing to availability of summary statistics only, and then applied incorrectly, it will bias the results. This is a well-described problem in regular fine-mapping (and by extension SNP-based colocalization), especially in meta-analyses of genome-wide association studies^37^. This is easily observed in our HBV results above where accuracy dropped significantly when simply using different LD panels. Things could become even more problematic if using HLA summary statistics from different ancestries, where difference in allele frequencies would lead SuSiE to assign high PIPs to entirely different alleles, even if using the correct LD reference panel. For example, in CKB, *HLA-DPB1*05:01* has a beta of 0.23 and a frequency of 37% (p = 2.1×10^-19^) while *HLA-DPB1*04:01* has a beta of −0.31 and a frequency of 37% (p = 2.5×10^-11^). In a Bangladeshi cohort^23^ using a related quantitative phenotype of opposite effect direction (level of Anti-HBs), *HLA-DPB1*05:01* has a beta of −1.03 and a frequency of 0.7% (p = 1.2×10^-5^) while *HLA-DPB1*04:01* has a beta of 0.49 and a frequency of 31% (p = 4.5×10^-30^). In both cohorts, *HLA-DPB1* is clearly associated (and likely causal) for HBV serological traits, but would lead to different PIPs due to differences in allele frequencies. Hence, like SNP-based colocalization, differences in genetic architecture across populations also prohibit the use of HLA-colocalization using two datasets from different ancestries.

In conclusion, HLA-colocalization is a new genetic causal inference method with good performance at the HLA. It requires few assumptions (essentially the same as for regular colocalization), is easy to implement with already existing tools, and performs well on simulated and real-world data. We believe it has the potential to advance the HLA field and lead to many clinical translational opportunities.

## Methods

### HLA-colocalization steps

The algorithm uses HLA allele association studies summary statistics and a population LD matrix as input. The alleles and LD architecture therefore need to be the same in both samples. It then works in two steps. First, we perform BVS using SuSiE and obtain PIPs or each allele. SuSiE is used because it provides an efficient way to approximate the posterior inclusion probabilities^38^. This step was done in R with the susie_rss function, with default parameters, and using all HLA alleles at the same time.

Second, to measure how similar each gene’s causal signature is, we perform Bayesian linear regression on each pair of PIPs. This is done using Stan^39^ in R, with the rstanarm package. We use the default priors used by rstanarm for linear regression. Specifically, the prior for the intercept term is Normal with a mean equal to the mean PIPs of the second trait, and a standard deviation of 2.5 times the standard deviation of the second trait. The prior for the slope is Normal with a mean of 0, and a standard deviation of 2.5 times the ratio of the standard deviation of the second trait and the standard deviation of the first trait. The probability of direction is then extracted for the slope coefficient (assuming that the coefficient is positive, otherwise colocalization is rejected). This regression step is done for each gene separately.

The final probability of HLA-colocalization is a function of the two steps. Specifically, there is colocalization if a gene has at least one pair of alleles with high PIPs in both traits, and if the slope of the regression is positive. The probability of each statement is then multiplied to give the following probability of colocalization (at each gene separately):

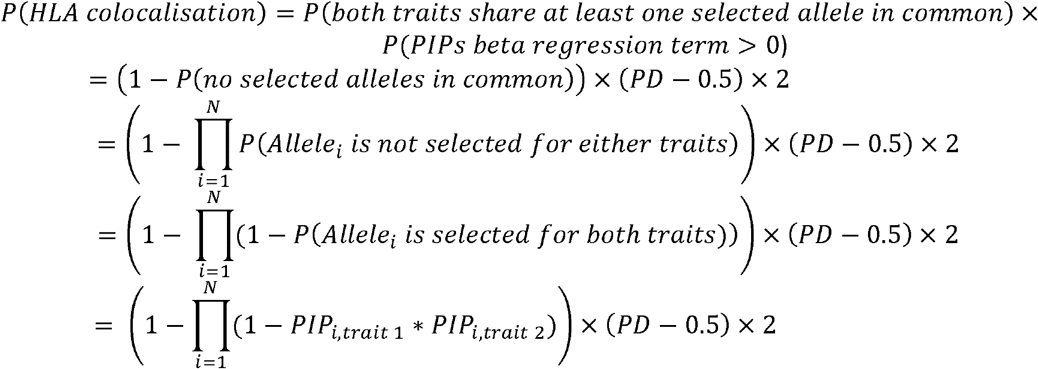

where N is the number of alleles at the HLA gene, *PIP*_*i*,*trait j*_ is the posterior inclusion probability of HLA allele i for trait j, and PD is the probability of direction of the Bayesian regression slope estimate at the HLA gene. The value (*PD* − 0.5) × 2 represents the size of the smallest credible interval around the Bayesian regression slope that overlaps the null. It approximates the probability that this slope is entirely contained within and infinity (1 above 0), to ensure that the method is robust to the choice of either trait 1 or trait 2 as the dependent variable in the regression. Note that in degenerate cases where the regression slope is below 0, we automatically set the probability of colocalization to 0.

Lastly the value in parentheses on the left-hand side of the bottom equation is related to the common SNP-based colocalization method of multiplying each PIP SNP by SNP and concluding that there is colocalization if at least one of the multiplication is high. Here, while this is not sufficient to claim HLA colocalization, however this formula clearly demonstrates that is still necessary.

### HLA allele data sources and association studies

For all UK Biobank analyses (including simulations, see section below), HLA alleles were obtained from previously published work^2^. Briefly, HLA alleles were called at a 3-field resolution using the HLA-HD algorithm^40^ on UK Biobank whole-exome sequences. For the HBV and liver disease analyses, HLA alleles were imputed at G-group resolution using whole-genome genotyping data and the Michigan Imputation Server multiethnic HLA imputation panel (v2)^41^. For the IMSGC multiple sclerosis analyses, HLA allele imputation was performed by the IMSGC, and is described elsewhere^42^.

Other than for the analysis from the IMSGC and the Taiwan Biobank (both described elsewhere^24,42^), all HLA association studies were performed using Regenie^43^ with an additive effect model (like genome-wide association studies). Age, sex, and the first 10 principal components were used as covariates. Approximate Firth regression penalty was used for case-control phenotypes using the default Regenie settings.

For the UK Biobank analyses, we also included recruitment center as a covariate, while geographical region was also used in CKB analyses. For EBV serologies, phenotypes were first inverse quantile normalized, then used as continuous variables. The HBV surface antigenemia is only reported as a binary trait in the CKB and was therefore analyzed as a case-control study. Multiple sclerosis was also analyzed as a categorical binary trait. For the binary traits in the UK Biobank, controls were selected as anybody who was not a case in the biobank. In CKB, controls were selected from the pre-specified control population, which adjusts for the by-design over-representation of patients with cancer and other chronic diseases in the cohort^8^.

### Simulation methods

To demonstrate the effectiveness of our method, we simulated two phenotypes with varying level of gene-level colocalization at the HLA. The simulation was done as follows. First, we assume that each HLA gene HLA-X has *N*_*X*_ alleles 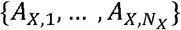. For the first phenotype (p1), we assign to each gene HLA-X a variance parameter 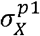, which represents the spread of the distribution of effects of each allele in that gene. Each allele *A*_*X*,*i*_ then has an associated effect on p1 distributed as 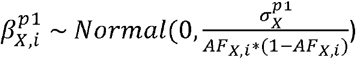, where *AF*_*X*,*i*_ is the allele frequency of the i^th^ allele of gene HLA-X. The reason for the denominator in the variance component of the normal distribution is to better reflect the fact that common variants have smaller effect sizes^44^. During the simulation we randomly set up to one third of 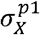to zero, denoting complete lack of causal effect of HLA-X on p1. We also randomly set up to all 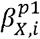 to zero, to denote complete lack of causal effect of allele *A*_*X*,*i*_ on p1. Lastly, we then center all 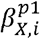 so that their allele frequency weighted average is 0. This represents the fact that the effect of an HLA allele at a gene is always expressed relative to the other alleles at that gene.

For the second phenotype (p2), every gene can be divided into two categories. First, if p1 and p2 do not colocalize at HLA-X, then we assign effects 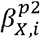 to each of its alleles in the same way that it was done for p1 above. Specifically, the simulation of 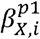 and 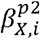 are totally independent. If p1 and p2 colocalize, then 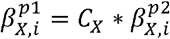, where *C*_*X*_ is a constant simulated independently for each gene. This is the same method used for SNP-based colocalization simulation^18^, and represents the fact that if two phenotypes share the same genetic determinants at an HLA gene, then alleles with a larger effect on the first phenotype should also have larger effect on the second. For each simulation, the number of causal genes for each phenotype was determined randomly (i.e. uniform distribution from 0 to the number of genes). From the number of causal genes for each gene, the number of shared causal gene was also determined randomly from a uniform distribution.

Using parameters above, we then simulate p1 and p2 for each participant, and add random noise to each simulation so that HLA genes explain on average 10% of the variance of the phenotypes. Lastly, HLA alleles association studies were performed on this simulated individual level data to obtain betas and standard errors. These were then used to perform HLA colocalization on the simulated data.

This was done in each of the 5 continental ancestry groups in the UK Biobank. For computational practicalities, the European ancestry group was limited to those who had serological measurements done (n = 8,158)^1,3^. Sample sizes were as follows for the 4 other groups: 8,725 participants of African genetic ancestry, 2,898 of Admixed American genetic ancestry, 2,647 of East Asian genetic Ancestry, and 9,449 of South Asian genetic ancestry.

We also performed a binary trait analysis. We used the same method as above to simulate betas on the liability scale, then transformed the results to binary phenotypes with the probit model. Note that due to decreased statistical power for binary traits, we simulated 10 times as many participants in this simulation as for the quantitative trait simulations above.

Lastly, we also ran a separate simulation with a number of single effect of 20, and obtained similar results (*Supp. Fig. 10-13*)

## Data Availability

All code necessary to perform HLA colocalization and the above simulation is available at https://github.com/DrGBL/hlacoloc. Primary data from the UKB and the CKB are available through their respective owners. All summary statistics needed to replicate our results are available on the git or on their respective publications when applicable.

## Ethics

All primary individual level participant data from the UKB was obtained using application 27449. The UKB has ethics approval from the North West Multi-centre Research Ethics Committee. Ethics approval for the CKB study was obtainedEthical Review Committee of the Chinese Centre for Disease Control and Prevention (Beijing, China, 005/2004) and the Oxford Tropical Research Ethics Committee, University of Oxford (UK, 025-04). Data from all other cohorts are publicly available summary statistics from their respective sources.

## Supplementary files

*Supplementary Data 1:* Colocalization results

*Supplementary Data 2:* Pathogen and autoimmune diseases colocalization full results

*Supplementary Figure 1:* Per ancestry ROC area under the curves for simulations of quantitative traits

*Supplementary Figure 2:* Characteristics of falsely colocalizing genes in our simulations

*Supplementary Figure 3:* HLA allele HLA-colocalization simulation results for binary traits

*Supplementary Figure 4:* Per ancestry ROC area under the curves for simulations of binary traits

*Supplementary Figure 5:* EBNA and multiple sclerosis HLA-colocalization in the UK Biobank

*Supplementary Figure 6:* VCA and multiple sclerosis HLA-colocalization in the IMSGC

*Supplementary Figure 7:* EBNA and multiple sclerosis HLA-colocalization in the IMSGC

*Supplementary Figure 8:* HBV and liver disease HLA-colocalization in the Taiwan Biobank

*Supplementary Figure 9:* Summary of pathogen and autoimmune diseases colocalizations

*Supplementary Figure 10:* HLA allele HLA-colocalization simulation results for quantitative traits with L=20

*Supplementary Figure 11:* Per ancestry ROC area under the curves for simulations of quantitative traits with L=20

*Supplementary Figure 12:* HLA allele HLA-colocalization simulation results for binary traits with L=20

*Supplementary Figure 13:* Per ancestry ROC area under the curves for simulations of binary traits with L=20

---

**Supplementary Figure 1:**
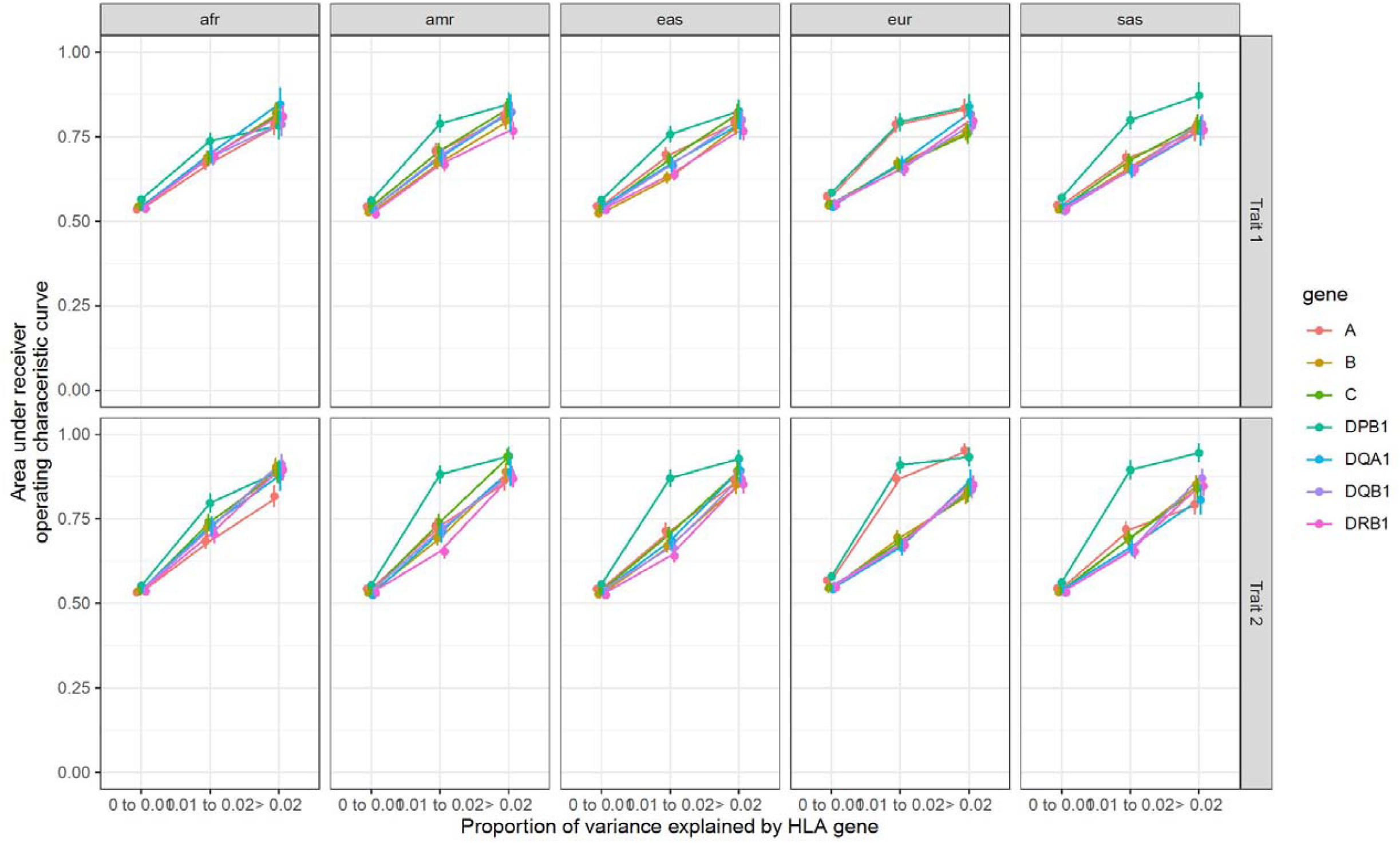
Per ancestry ROC area under the curves for simulations of quantitative traits. Area under the ROC curves of HLA-colocalization PIPs for different variance explained per gene and genetic ancestries for the simulation of the quantitative traits. Legend: afr: African genetic ancestry, amr: Admixed American genetic ancestry, eas: East Asian genetic ancestry, eur: European genetic ancestry, sas: South Asian genetic ancestry.

**Supplementary Figure 2:**
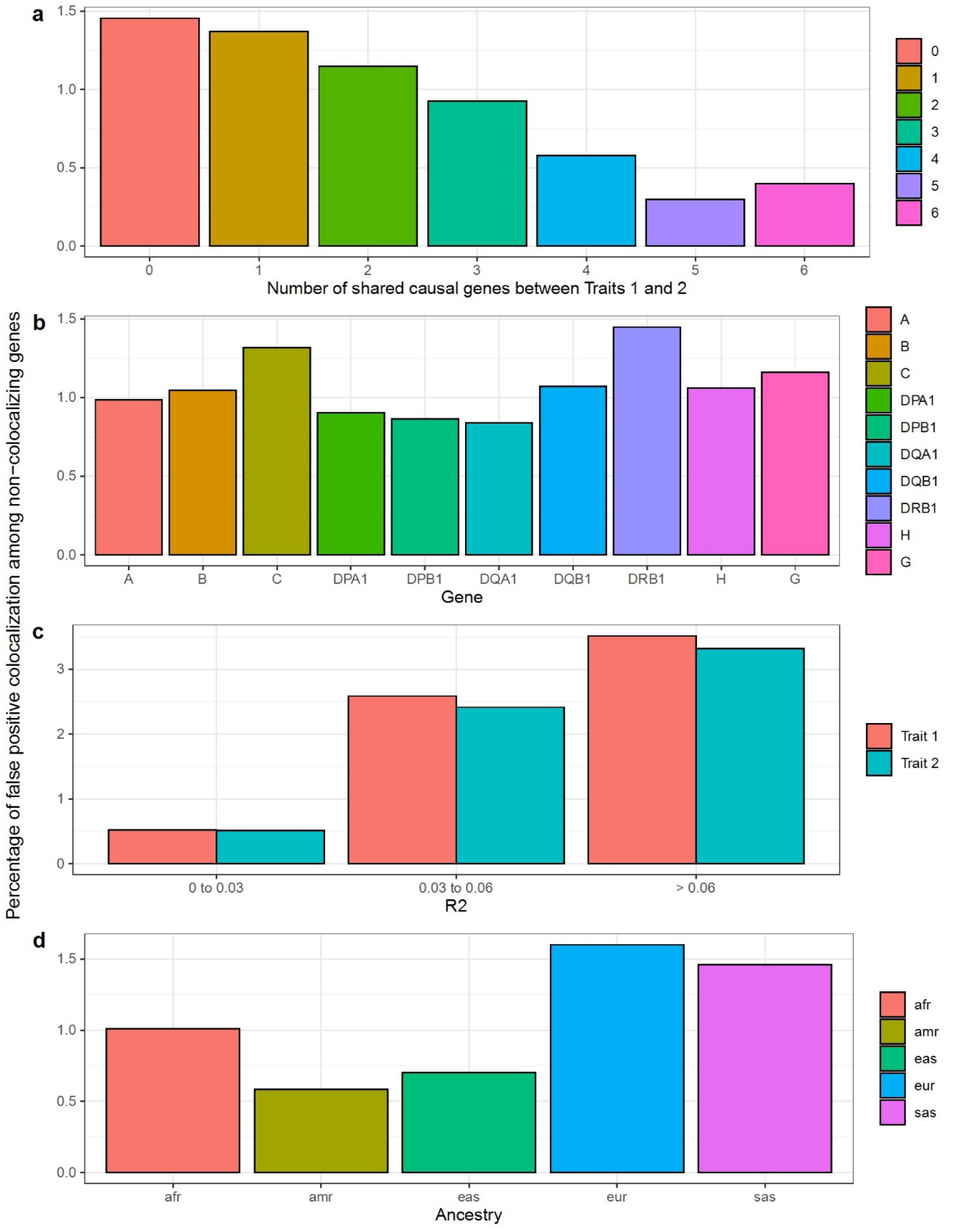
False colocalizing genes. We calculated the proportion of non-colocalizing genes with a probability of colocalization above 80% under varying circumstances. **a)** The proportion was higher in simulations where less genes were truly colocalizing then in simulations with more, since more truly colocalizing genes decreases the chance of a random false colocalization. **b)** The proportion was higher at HLA-DRB1 (1.45%) and HLA-C (1.32%) than other genes (e.g. HLA-DQA1 0.84%). **c)** The proportion was higher in gene which explained a larger portion of the traits, since genes with lower R2 are less likely to colocalize. d) The proportion was slightly higher in Europeans (1.6%) compared to other ancestries (e.g. admixed Americans at 0.58%). Overall, only 1.1% of non-colocalizing genes were given a probability of colocalization above 80%.

**Supplementary Figure 3:**
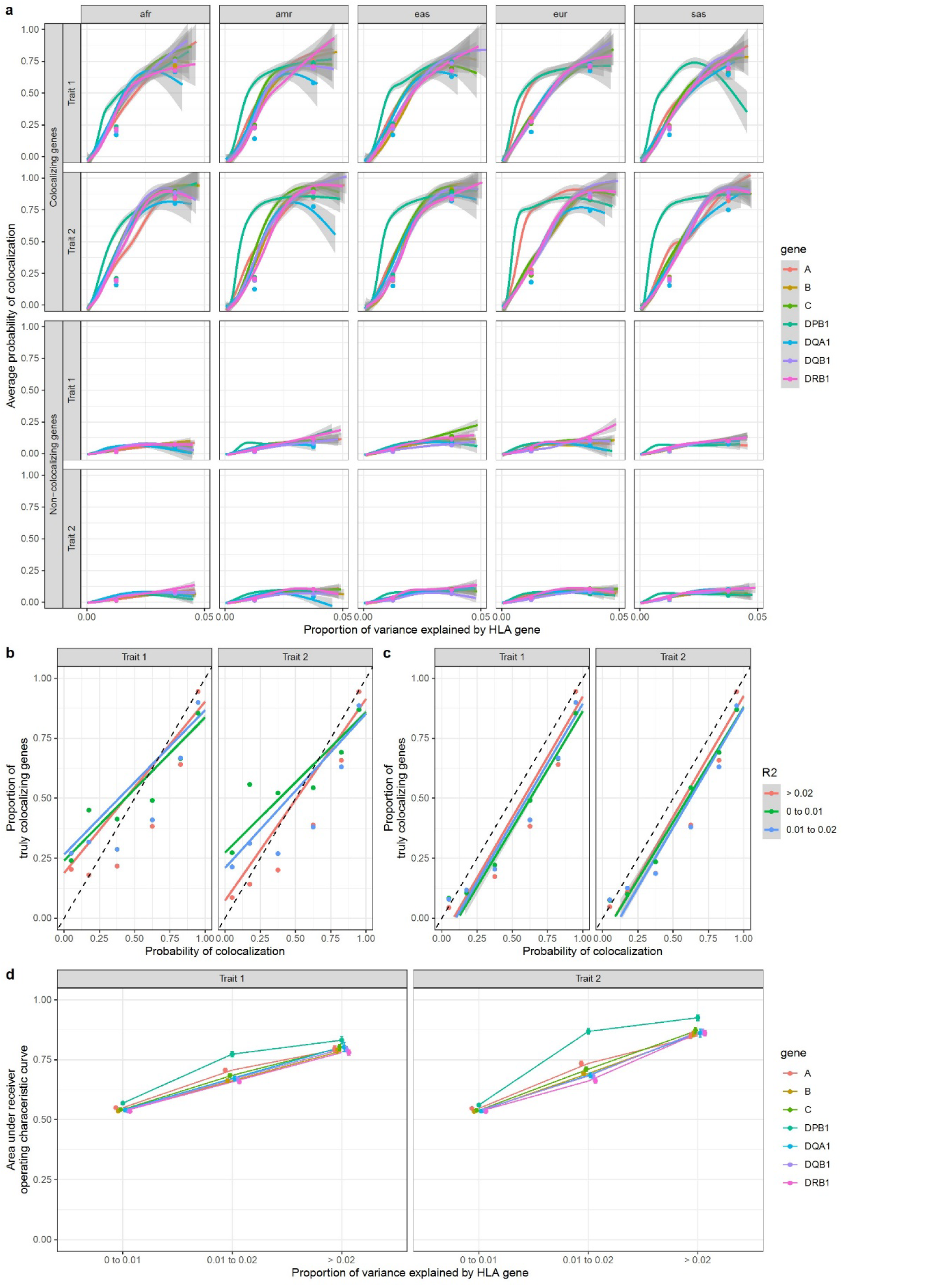
HLA allele HLA-colocalization simulation results for binary traits. Pairs of quantitative traits were simulated having either true overlap, or no true overlap between causal HLA alleles, using a bivariate normal model as described in Methods. In each simulation a total proportion of trait variance explained was assumed. A total of 50,000 simulations (10,000 per ancestry group) were performed covering different parameter values (Methods). HLA allele distributions were simulated using UK Biobank participants. **a)** The average posterior probability of colocalization in truly colocalizing increases with the amount of phenotype variance explained by each gene, as expected. The average posterior probability of colocalization in truly non-colocalizing genes remains stable with increasing variance explained. The lines were drawn using a generalized additive model with geom_smooth in R. The grey area represents 95% confidence intervals. The individual dots represent the average in the corresponding variance bins. **b)** The proportion of simulated genes that were truly colocalizing shown as a function of the probability of colocalization. This is close to the identity line, though errs on the more conservative side for genes with lower R2. **c)** The deviation from the identity line is largely due to situations where SuSiE is unable to assign a PIP larger than 50% in at least one allele at a gene. When we restrict to genes with a minimal PIP of 50%, the method is almost perfectly calibrated. **d)** Average area under the curve as a function of variance explained for each gene. For this plot, average ROC area under the curve across ancestry was shown. Legend: afr: African genetic ancestry, amr: Admixed American genetic ancestry, eas: East Asian genetic ancestry, eur: European genetic ancestry, sas: South Asian genetic ancestry.

**Supplementary Figure 4:**
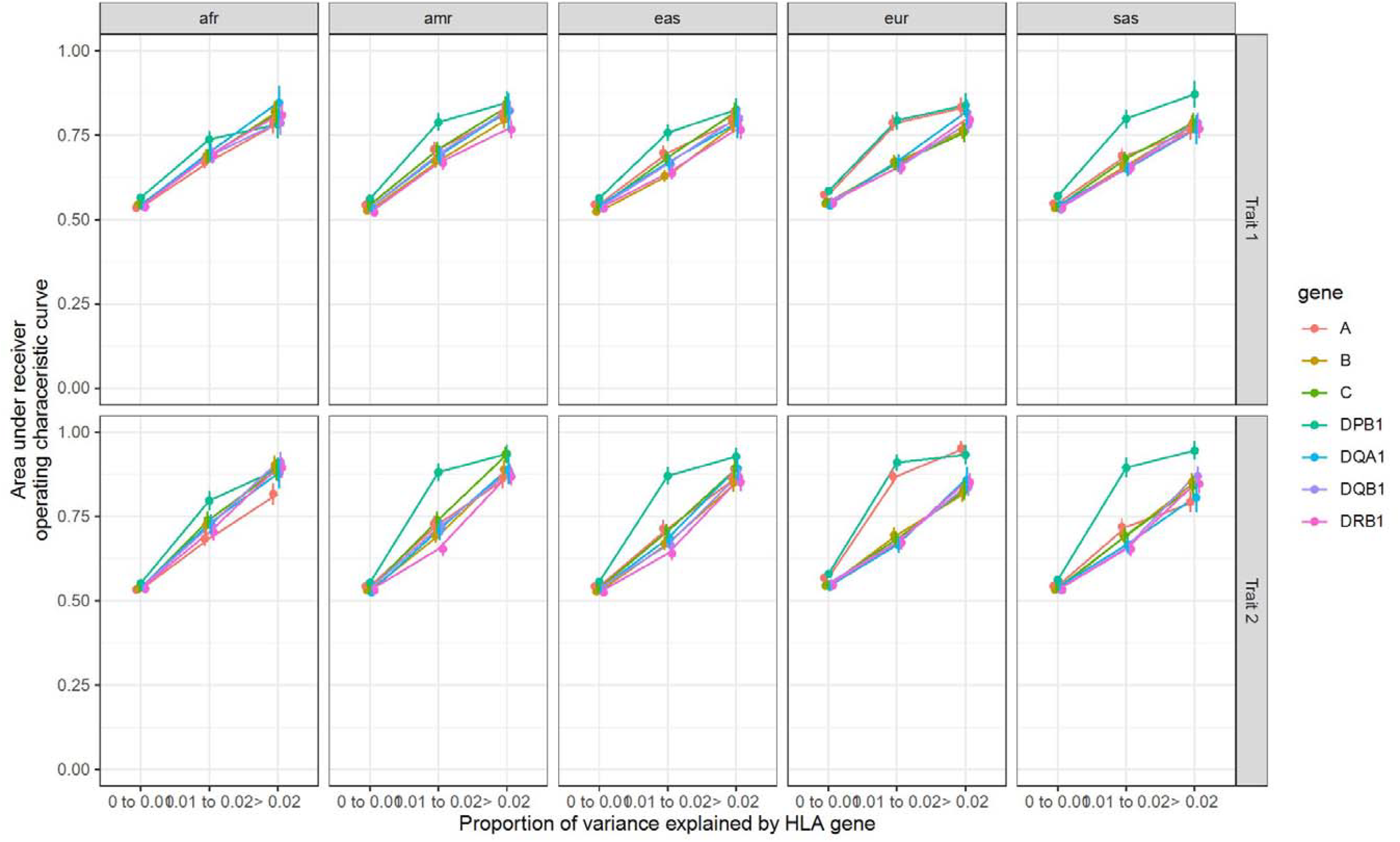
Per ancestry ROC area under the curves for simulations of binary traits. Area under the ROC curves of HLA-colocalization PIPs for different variance explained per gene and genetic ancestries for the simulation of the binary traits. Legend: afr: African genetic ancestry, amr: Admixed American genetic ancestry, eas: East Asian genetic ancestry, eur: European genetic ancestry, sas: South Asian genetic ancestry.

**Supplementary Figure 5:**
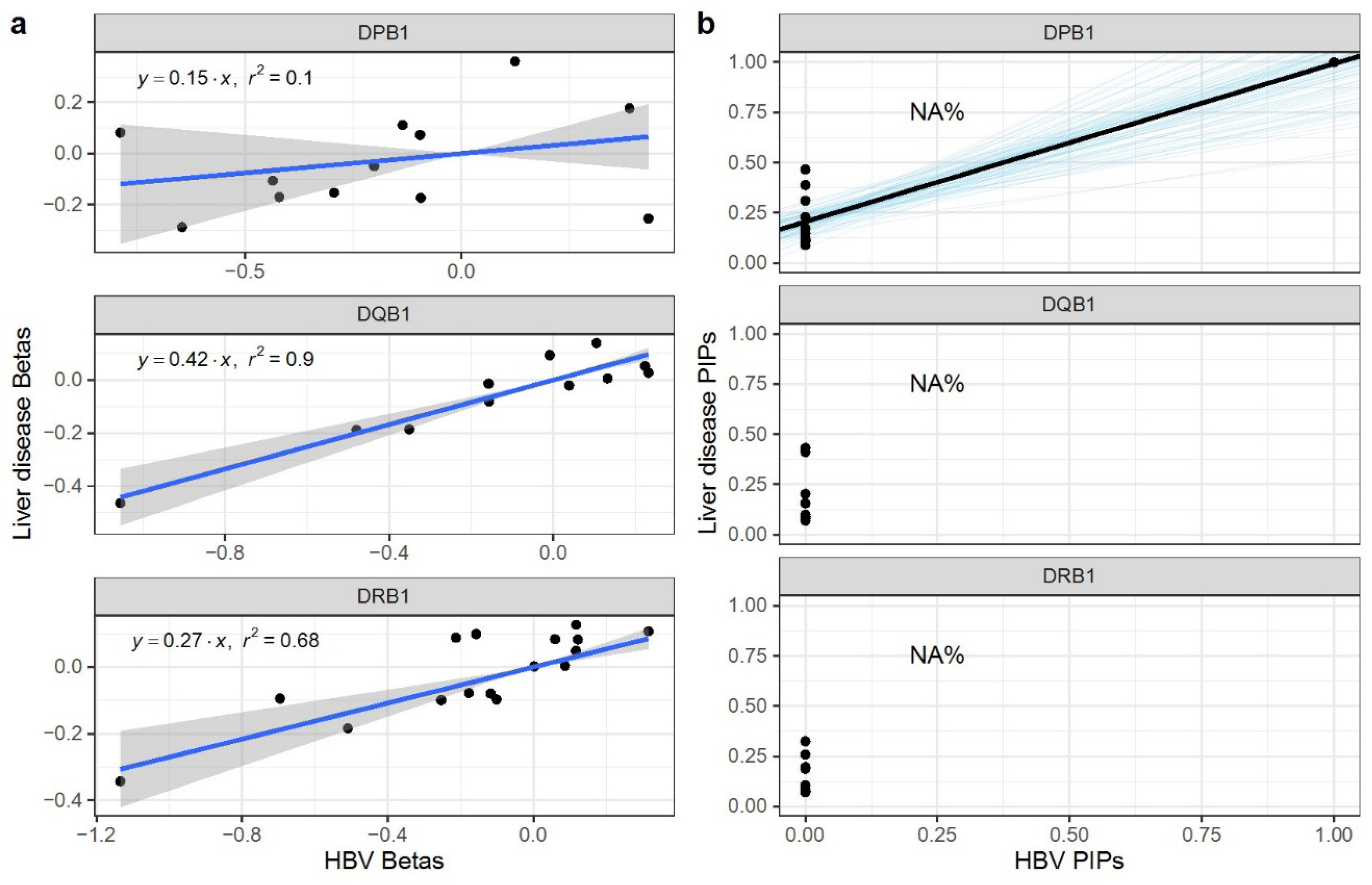
Hepatitis B (HBV) and liver disease HLA-colocalization in the Taiwan Biobank. **a)** linear regression (with 95% confidence intervals) of beta coefficients from the additive HLA allele association studies. **b)** Bayesian regression of liver disease PIPs on HBV PIPs causal signatures. The black lines show the regression fit, while the blue lines show 100 random draws from the posterior distributions. The resulting probabilities of HLA-colocalization (P_coloc) are also written for ease. Once again, we observe HLA-colocalization at *HLA-DPB1*.

**Supplementary Figure 6:**
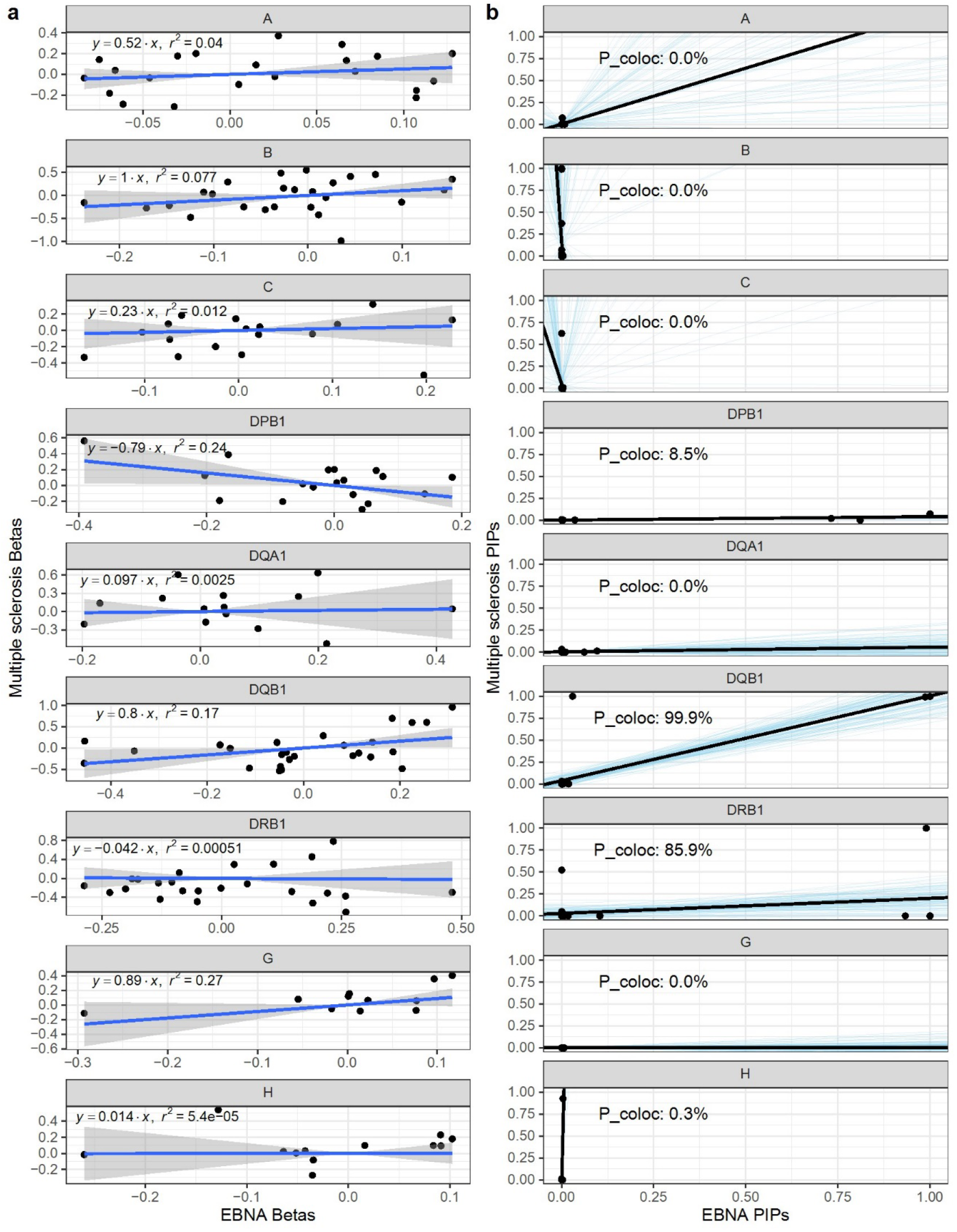
EBNA and multiple sclerosis HLA-colocalization in the UK Biobank. **a)** linear regression (with 95% confidence intervals) of beta coefficients from the additive HLA allele association studies. **b)** Bayesian regression of multiple sclerosis PIPs on EBNA PIP causal signatures. The black lines show the regression fit, while the blue lines show 100 random draws from the posterior distributions. The resulting probabilities of HLA-colocalization (P_coloc) are also written for ease.

**Supplementary Figure 7:**
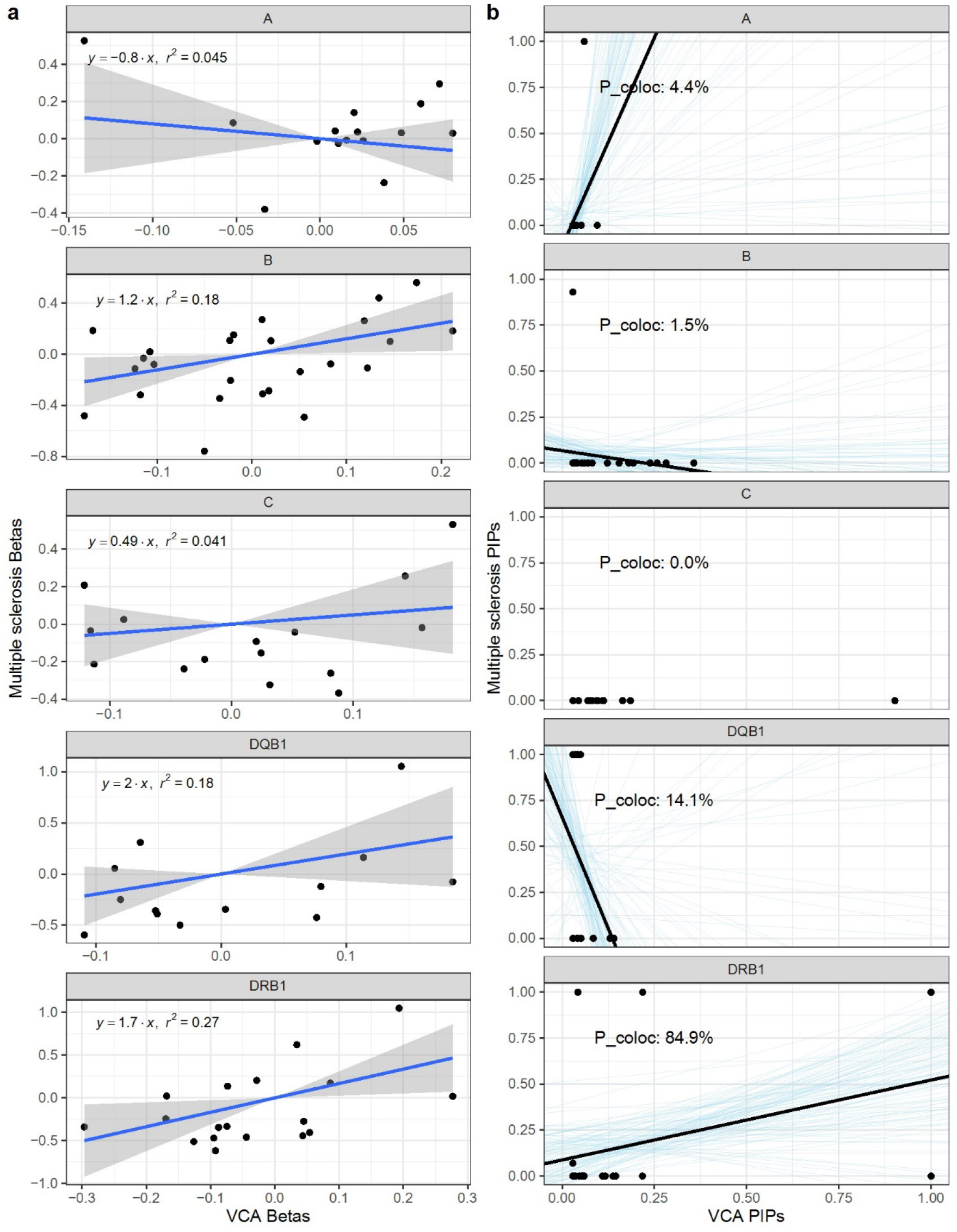
VCA and multiple sclerosis HLA-colocalization in the IMSGC. **a)** linear regression (with 95% confidence intervals) of beta coefficients from the additive HLA allele association studies. **b)** Bayesian regression of multiple sclerosis PIPs on VCA PIP causal signatures. The black lines show the regression fit, while the blue lines show 100 random draws from the posterior distributions. The resulting probabilities of HLA-colocalization (P_coloc) are also written for ease.

**Supplementary Figure 8:**
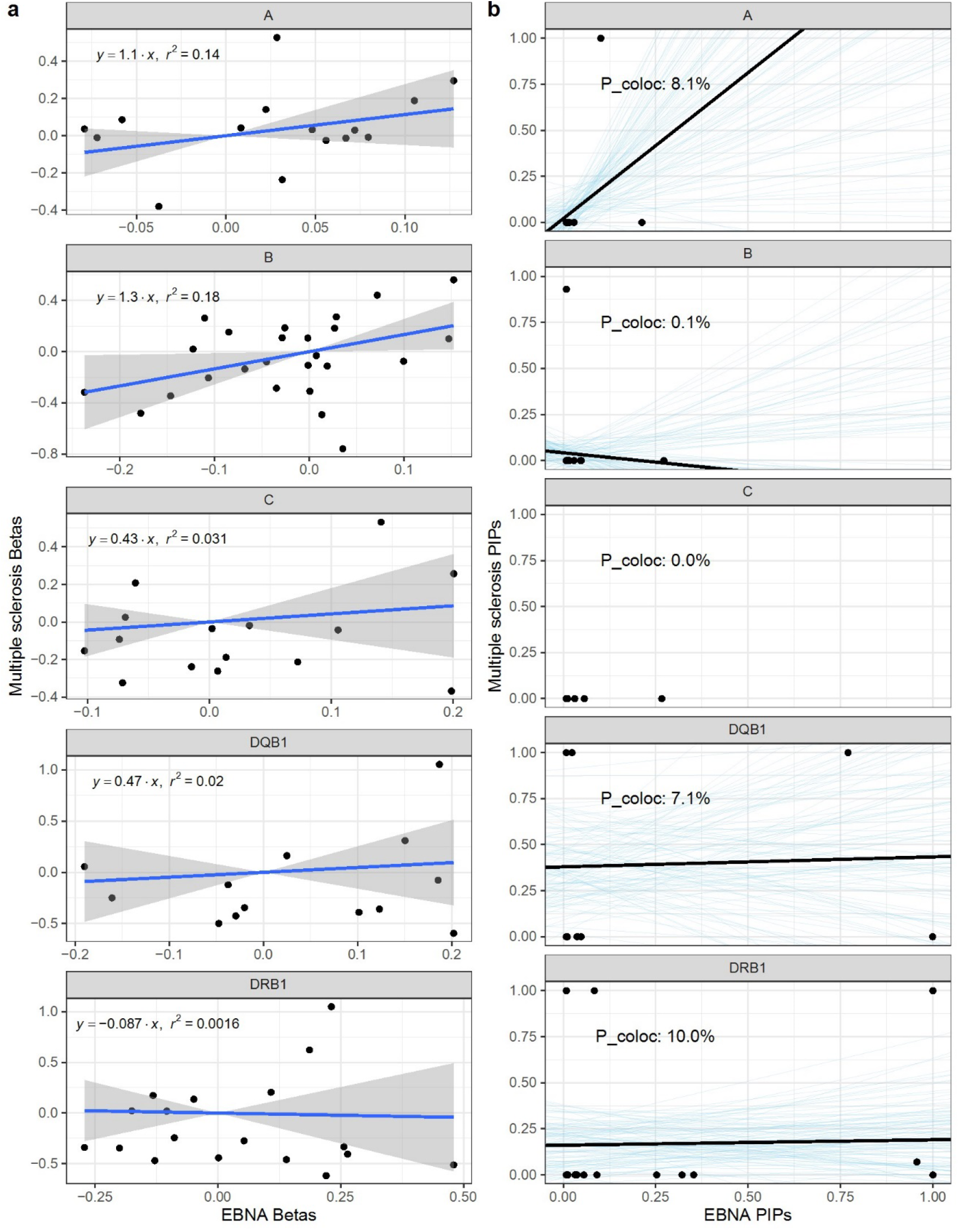
EBNA and multiple sclerosis HLA-colocalization in the IMSGC. **a)** linear regression (with 95% confidence intervals) of beta coefficients from the additive HLA allele association studies. **b)** Bayesian regression of multiple sclerosis PIPs on EBNA PIP causal signatures. The black lines show the regression fit, while the blue lines show 100 random draws from the posterior distributions. The resulting probabilities of HLA-colocalization (P_coloc) are also written for ease.

**Supplementary Figure 9:**
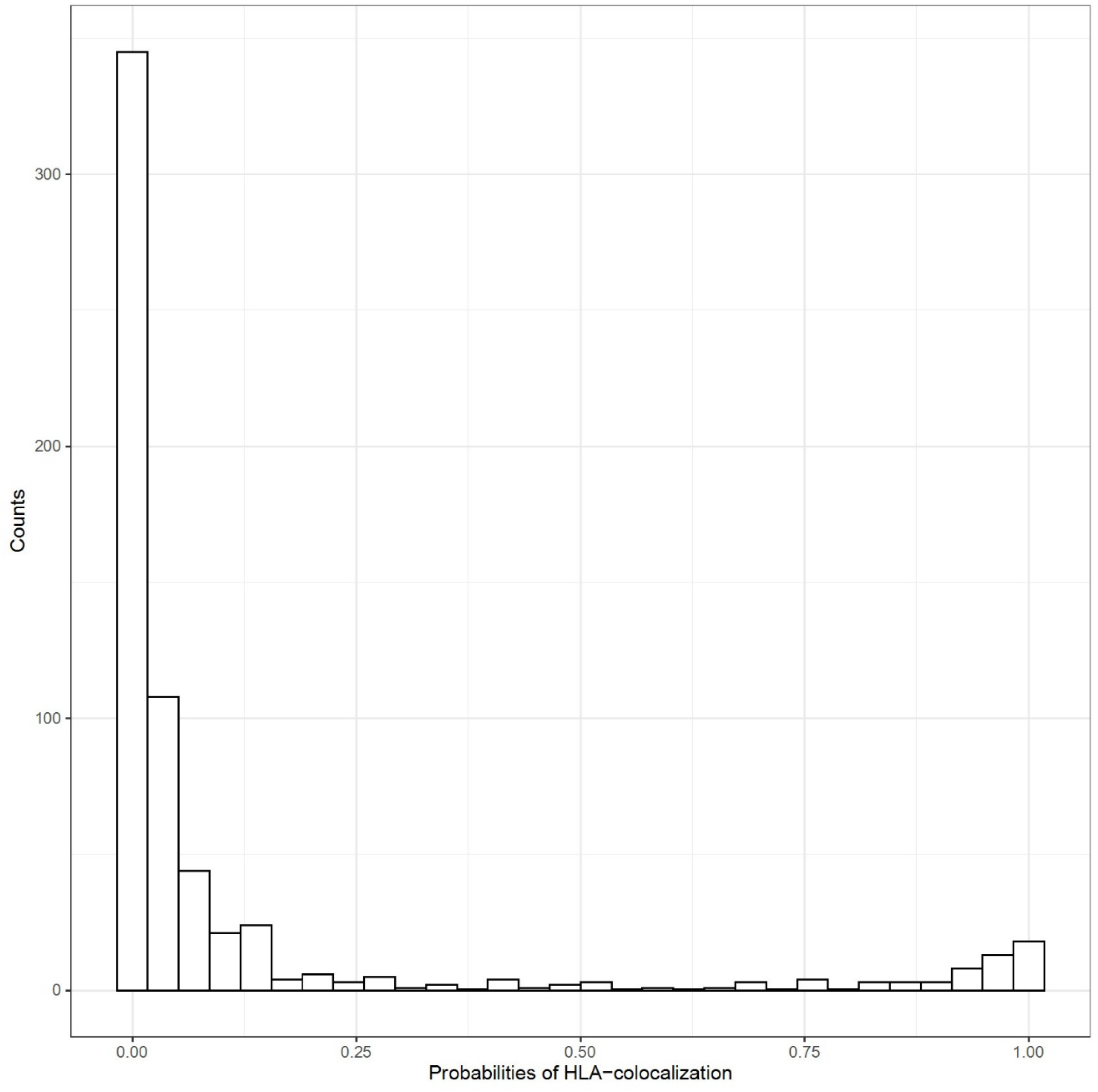
pathogen and auto-immune traits colocalization results. Distribution of HLA-colocalization probabilities for all pairs of pathogen serology and auto-immune diseases traits (n = 630 pathogen to autoimmune diseases pairs). As can be seen, most pairs of traits do not colocalize, which is expected and suggest that our method is well calibrated to complex real-world data. See Supp. Data 2 for the full results.

**Figure S10.**
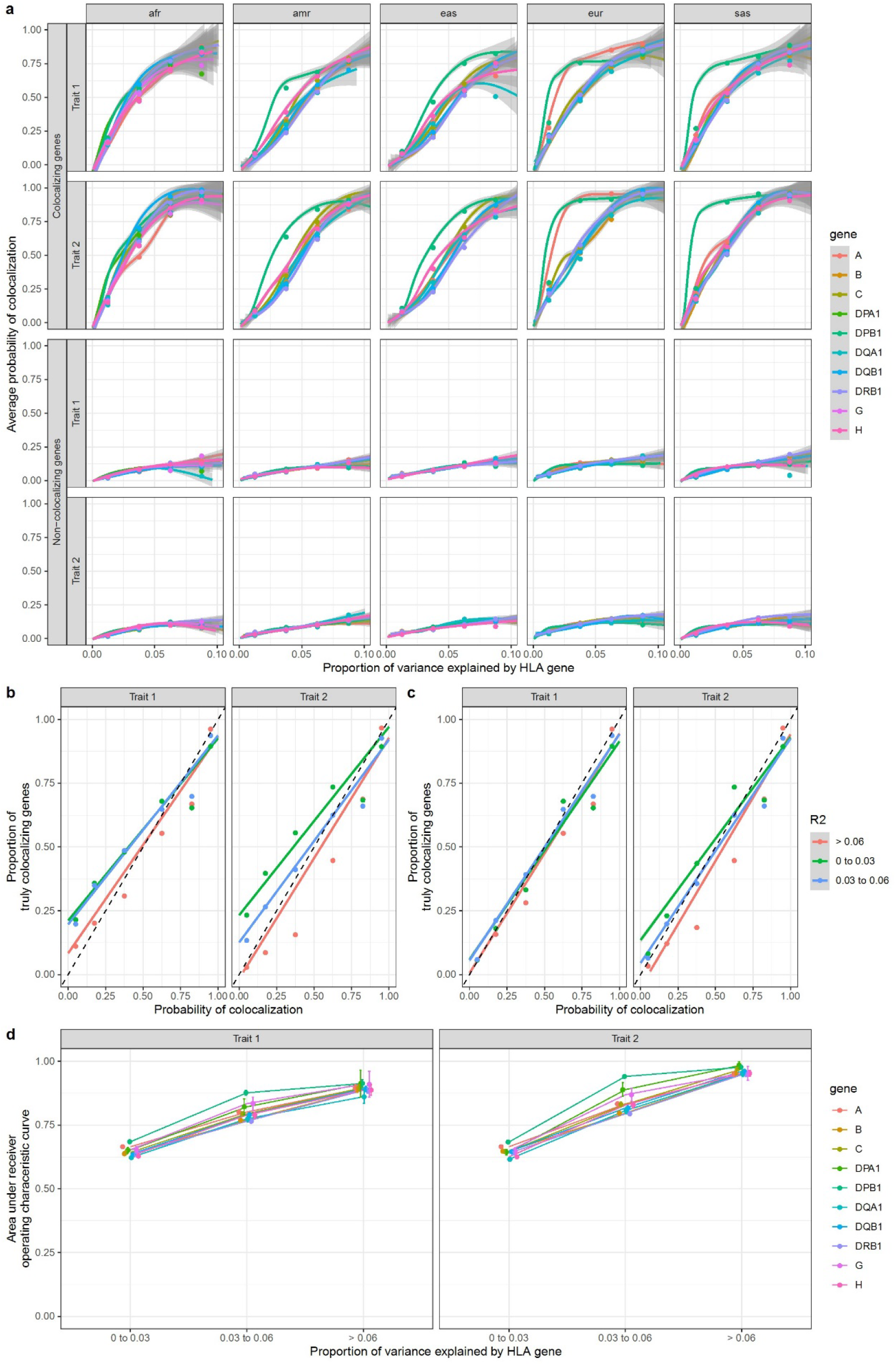
HLA allele HLA-colocalization simulation results for quantitative traits with L = 20. This simulation was done using the number of single effect option of SuSiE at 20 i.e. (L = 20). **a)** The average posterior probability of colocalization in truly colocalizing increases with the amount of phenotype variance explained by each gene, as expected. The average posterior probability of colocalization in truly non-colocalizing genes remains stable with increasing variance explained. The lines were drawn using a generalized additive model with geom_smooth in R. The grey area represents 95% confidence intervals. The individual dots represent the average in the corresponding variance bins. **b)** The proportion of simulated genes that were truly colocalizing shown as a function of the probability of colocalization. This is close to the identity line, though errs on the more conservative side for genes with lower R2. **c)** The deviation from the identity line is largely due to situations where SuSiE is unable to assign a PIP larger than 50% in at least one allele at a gene. When we restrict to genes with a minimal PIP of 50%, the method is almost perfectly calibrated. **d)** Average area under the curve as a function of variance explained for each gene. For this plot, average ROC area under the curve across ancestry was shown. Legend: afr: African genetic ancestry, amr: Admixed American genetic ancestry, eas: East Asian genetic ancestry, eur: European genetic ancestry, sas: South Asian genetic ancestry.

**Supplementary Figure 11:**
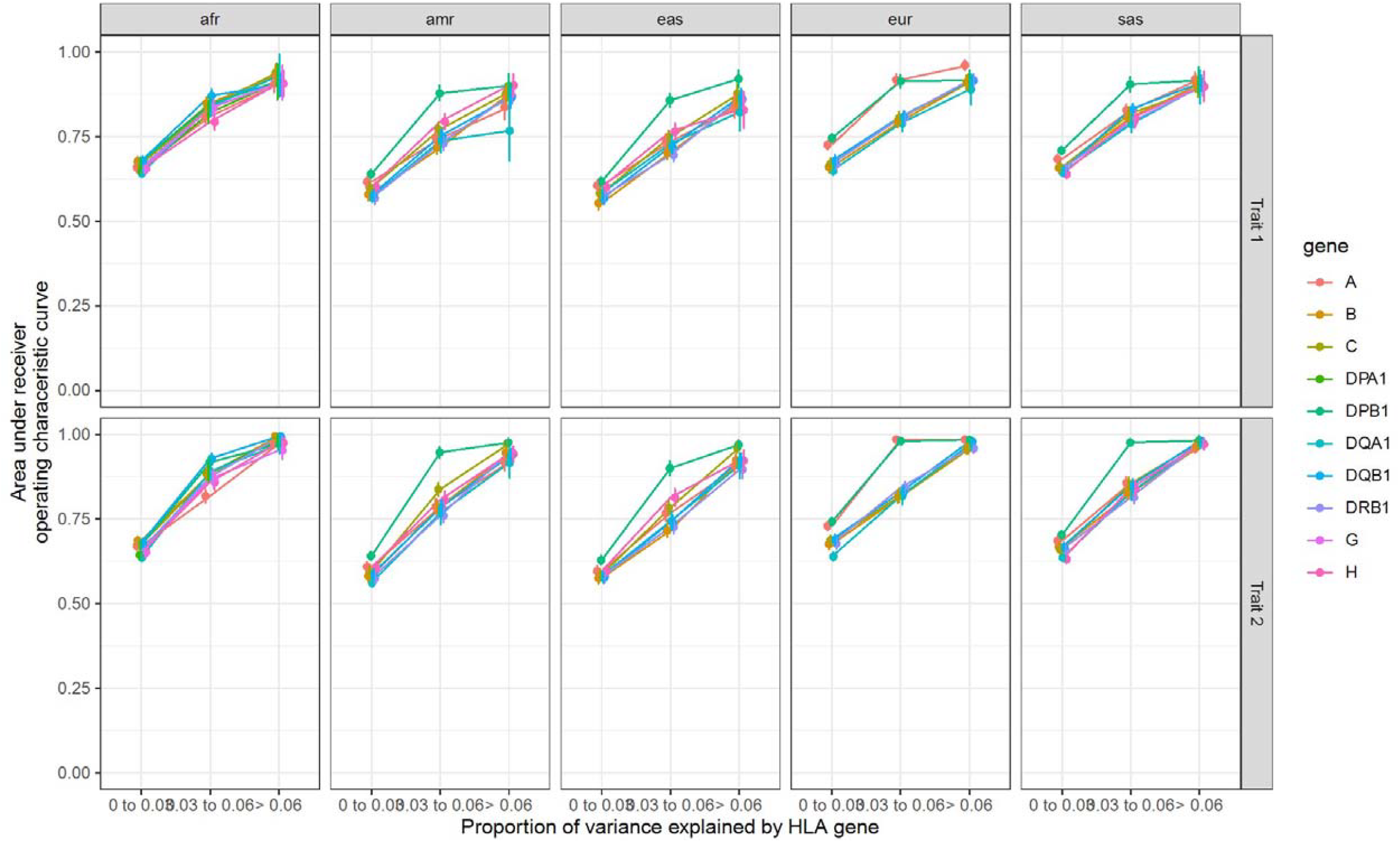
Per ancestry ROC area under the curves for simulations of quantitative traits with L = 20. This simulation was done using the number of single effect option of SuSiE at 20 i.e. (L = 20). Area under the ROC curves of HLA-colocalization PIPs for different variance explained per gene and genetic ancestries for the simulation of the quantitative traits. Legend: afr: African genetic ancestry, amr: Admixed American genetic ancestry, eas: East Asian genetic ancestry, eur: European genetic ancestry, sas: South Asian genetic ancestry.

**Supplementary Figure 12:**
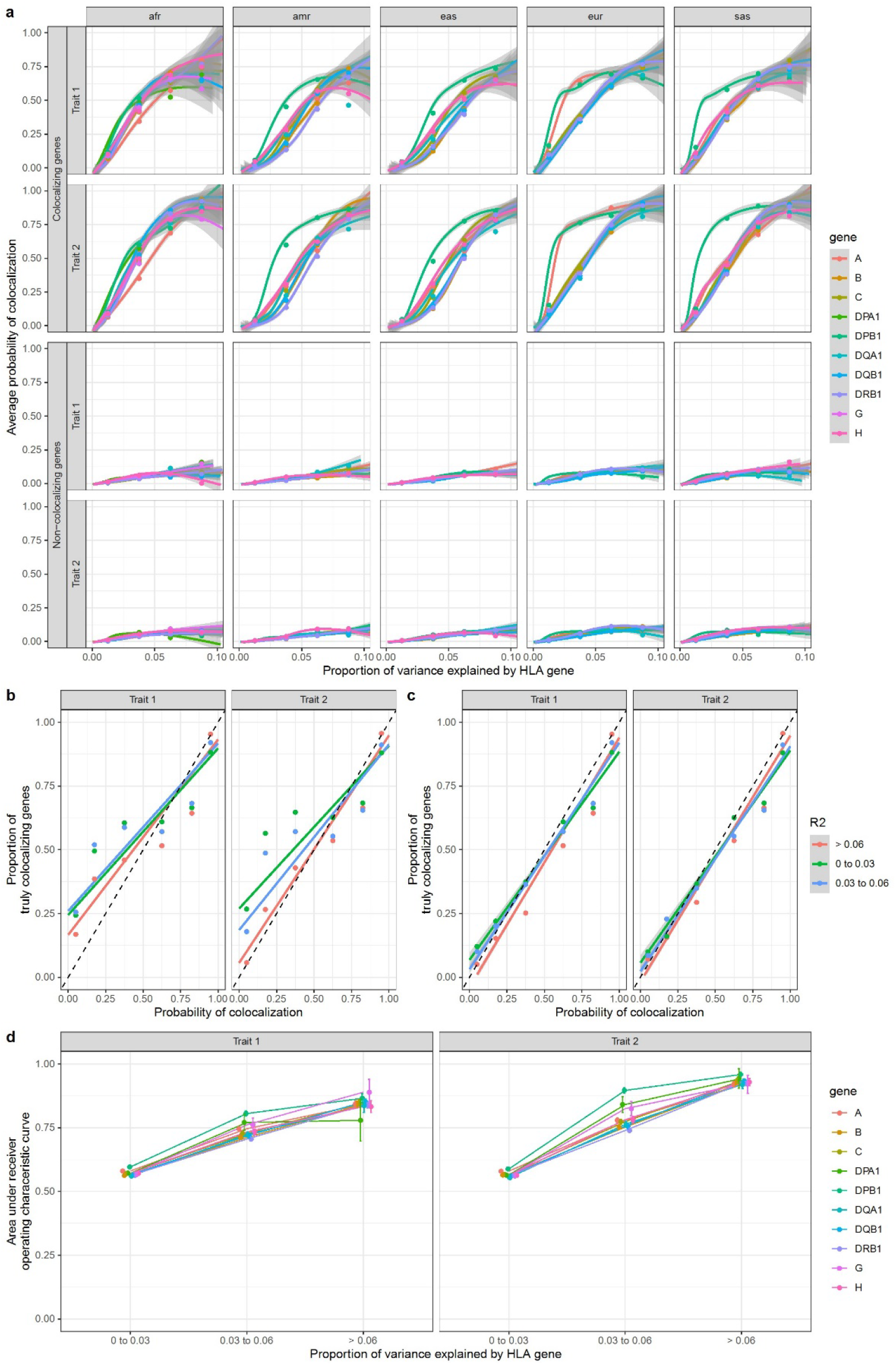
HLA allele HLA-colocalization simulation results for binary traits with L = 20. This simulation was done using the number of single effect option of SuSiE at 20 i.e. (L = 20). **a)** The average posterior probability of colocalization in truly colocalizing increases with the amount of phenotype variance explained by each gene, as expected. The average posterior probability of colocalization in truly non-colocalizing genes remains stable with increasing variance explained. The lines were drawn using a generalized additive model with geom_smooth in R. The grey area represents 95% confidence intervals. The individual dots represent the average in the corresponding variance bins. **b)** The proportion of simulated genes that were truly colocalizing shown as a function of the probability of colocalization. This is close to the identity line, though errs on the more conservative side for genes with lower R2. **c)** The deviation from the identity line is largely due to situations where SuSiE is unable to assign a PIP larger than 50% in at least one allele at a gene. When we restrict to genes with a minimal PIP of 50%, the method is almost perfectly calibrated. **d)** Average area under the curve as a function of variance explained for each gene. For this plot, average ROC area under the curve across ancestry was shown. Legend: afr: African genetic ancestry, amr: Admixed American genetic ancestry, eas: East Asian genetic ancestry, eur: European genetic ancestry, sas: South Asian genetic ancestry.

**Supplementary Figure 13:**
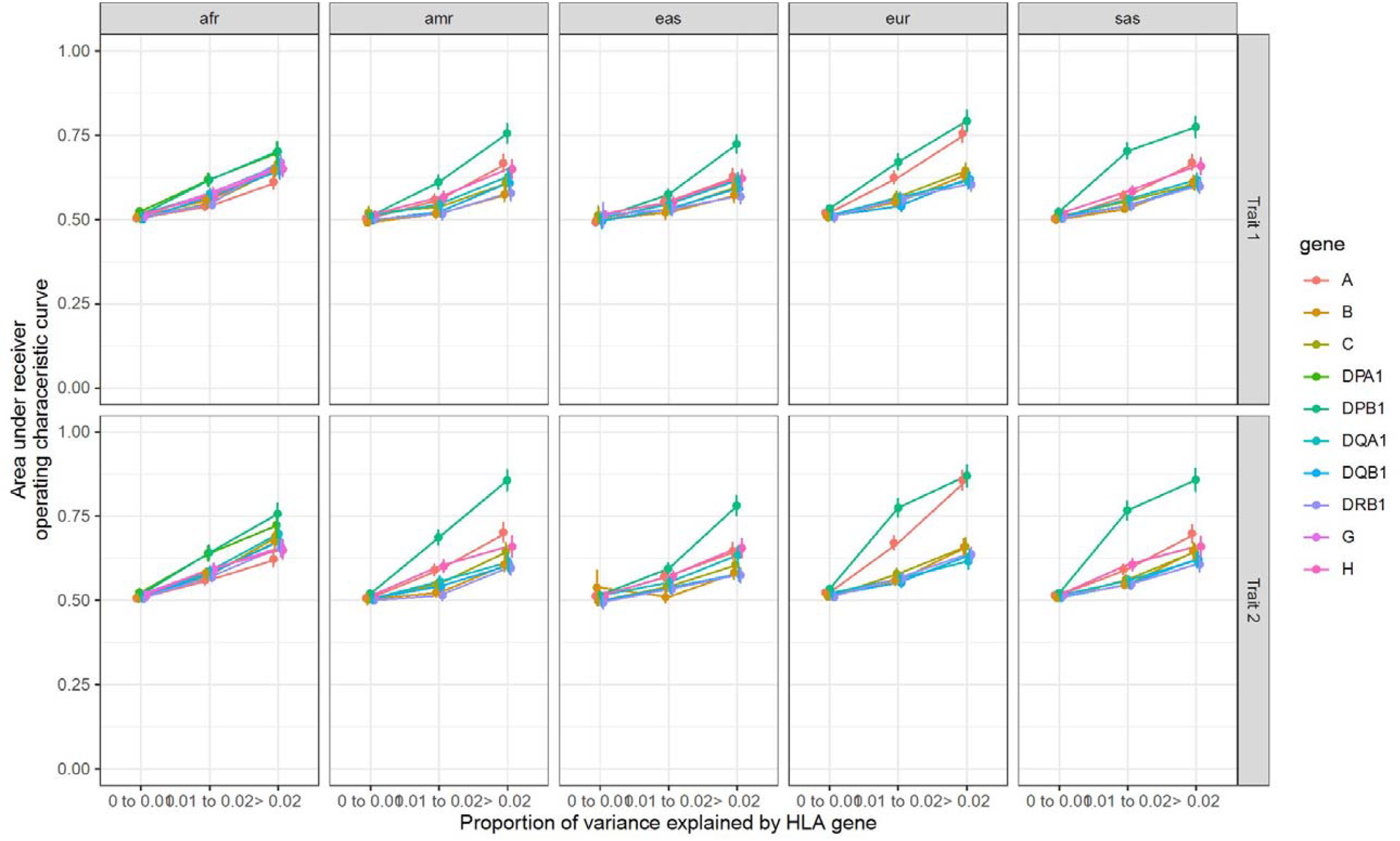
Per ancestry ROC area under the curves for simulations of binary traits with L = 20. This simulation was done using the number of single effect option of SuSiE at 20 i.e. (L = 20). Area under the ROC curves of HLA-colocalization PIPs for different variance explained per gene and genetic ancestries for the simulation of the binary traits. Legend: afr: African genetic ancestry, amr: Admixed American genetic ancestry, eas: East Asian genetic ancestry, eur: European genetic ancestry, sas: South Asian genetic ancestry

